# Modelling Acral Melanoma in Admixed Brazilians Uncovers Genomic Drivers and Targetable Pathways

**DOI:** 10.1101/2025.08.08.25332963

**Authors:** Annie Cristhine Moraes Sousa-Squiavinato, Sara Santos Bernardes, Flávia C. Aguiar, Antonio C. Facciolo, Martín del Castillo Velasco Herrera, J. Rene C. Wong-Ramirez, Patricia Basurto-Lozada, Aretha Brito Nobre, Geethanjali Annamalai, Rebecca Martins Cadimo do Nascimento, Jacqueline Boccacino, Rafaela Fagundes, Pedro Sodré do R. Barros, Mariana de Moraes Pitombo, Rebeca Olvera-León, Larissa Satiko Alcantara Sekimoto Matsuyama, Jamie Billington, Ian Vermes, Irving Simonin-Wilmer, Danielle G. Carvalho, João Pedro Cavalcante Simoes, Priscila Valverde Fernandes, Luiz Fernando Nunes, Andreia Cristina de Melo, Jadivan Leite de Oliveira, Meenhard Herlyn, Andrew E. Aplin, Carla Daniela Robles-Espinoza, David J. Adams, Patricia A. Possik

## Abstract

Acral melanoma (AM) is an aggressive melanoma subtype with limited therapeutic options and poor outcomes. In non-European descent and admixed populations, like those residing in Latin America, AM accounts for a significant proportion of cutaneous melanoma cases. Here, we performed comprehensive genomic and functional profiling of AM from a uniquely diverse Brazilian cohort. Whole-exome and transcriptome sequencing revealed low mutation burden and predominance of copy number alterations, including high-amplitude focal amplifications termed hailstorms. These hailstorms frequently affected chromosomes 11, 5 and 22 and key oncogenes such as *CCND1*, *GAB2*, *CDK4*, and *TERT*. The presence of hailstorms in the long arms of chromosomes 11 and 22 was associated with higher focal copy number burden and loss of DNA damage response genes (*ATM*, *CHEK1*), suggesting a permissive genomic environment driving structural instability. To explore the unique genomic context of AM, we established a comprehensive collection of patient-derived xenograft (AM-PDX) models that faithfully retain the histopathological and genomic features of the original tumours. Functional exploration of AM-specific vulnerabilities through pharmacological and CRISPR/Cas9 knockout screenings identified strong sensitivity to targeting MAPK, CDK4/6, MDM2, and WEE1 pathways. Notably, the pan-RAS(ON) inhibitor RMC-7977 effectively reduced viability in *NRAS*-, *KRAS*-, and *KIT*-mutant AM cell lines. Finally, CRISPR screens revealed dependencies selectively essential in AM, including *CRKL* and *SF3B4*, highlighting previously unrecognized vulnerabilities. Our findings emphasize the distinct biology of AM compared to other subtypes of melanoma, provide a valuable resource of models reflective of Latin American ancestry, and identify potential drivers and therapeutic targets.

## INTRODUCTION

Acral melanoma (AM) affects the glabrous skin of the soles and palms and subungual regions. The prognosis of AM is poor due to its aggressive phenotype, typically with high rates of ulcerated lesions and regional metastases, as well as late diagnosis, high recurrence rates, and the challenges of surgical treatment of functionally important anatomical sites^1,2^. Systemic treatment options are limited, with fewer patients benefiting from targeted therapies and generally poorer responses to immunotherapy, contributing to worse overall survival compared to advanced non-acral cutaneous melanoma (CM) ^3–6^.

Genomically, AM exhibits a lower frequency of well-characterized driver mutations that are commonly observed in ultraviolet radiation (UVR)-associated melanoma, such as *BRAF*^V600E^ mutations, which occur in sun-exposed body sites such as the face and trunk. In addition, while AM is characterized by fewer point mutations and small insertions and deletions, it shows a higher frequency of complex structural alterations, including amplifications and chromosomal aberrations^7–13^.

Compared to the other melanoma subtypes, AM remains understudied. This is partially attributed to the low proportion of cases among European descent populations in the Global North, where most genomic, preclinical and clinical research, as well as drug development efforts, has historically been centered^14,15^. In Western European countries, AM constitutes 2-3% of all melanoma cases^15^. In contrast, this proportion is higher in Asian countries, representing about 40% of melanomas in Korea, 50% in Singapore, and 58% in Taiwan^16–18^. In South Africa, AM proportion varies by province, ranging from 5 to 20%^19,20^. In Latin America, AM holds considerable epidemiological importance^21–23^ and the variation in AM as a proportion of melanoma cases likely reflects population diversity. For instance, it is notably higher in countries with high Amerindian ancestry, such as Mexico (44%)^24^ and Peru (32%)^25^, compared to Argentina (3.3%), where the population is predominantly of European descent^26^.

In Brazil, AM accounts for 10-15% of all CM cases^27,28^. Between 1997 and 2014, 3878 melanoma cases were diagnosed at the Brazilian National Cancer Institute in Rio de Janeiro (Instituto Nacional de Câncer, INCA), a tertiary referral centre under the Ministry of Health, which provides public health care to cancer patients. Five hundred and twenty-nine of these cases were diagnosed as AM, most of which were ulcerated (79%) and thick at diagnosis (median Breslow thickness: 5.0 mm). Additionally, 81% of patients had low socioeconomic status, measured by years of schooling, which was linked to worse outcomes in univariate analysis^28^. A closer look at stage I and II AM patients revealed that 28% experienced disease recurrence within five years, with half presenting with ulcerated tumours and a median Breslow thickness of 4.4 mm, consistent with the late diagnosis of AM worldwide^29^.

In addition to the need to increase public health efforts focused on accurate detection and prevention, the limited understanding of predisposing factors, causative agents, and molecular alterations results in a lack of reliable markers for early diagnosis and prognosis and effective treatments for AM patients. Notably, while it is known that the genetic alterations in AM differ substantially from UVR-related melanoma, therapeutic strategies tailored specifically for AM patients are unavailable, representing a significant unmet need in terms of clinical care.

Various experimental cancer models have been proposed as tools to identify specific biomarkers and therapeutic targets. Among those, patient-derived xenograft (PDX) models have been proven especially valuable as they preserve the cellular and histopathological structure of the original tumour, as well as its genomic features^30–32^. While PDX platforms have been extensively used in preclinical melanoma studies, most studies have focused on UV-associated melanoma from Europeans, or melanomas from Asian descent populations, with limited/no exploration of AM from Latin America^33–36^.

In this study, we assembled and analysed an extensive collection of AM samples and models derived from patients treated at INCA. Although *KRAS/NRAS/HRAS* were the most frequently mutated genes, most tumours were wild type for these genes and other common drivers such as *BRAF*, *NF1*, or *KIT*. Amplifications were common, affecting a significant portion of the tumour genome and often characterized by the presence of hailstorms, most frequently observed on chromosome 11. Capturing these features, we created AM-PDX models which faithfully replicated the key histopathological and genetic characteristics of patient tumours. By employing unbiased CRISPR/Cas9 knockout and pharmacological screens, we demonstrate that these models can be used to uncover AM-selective vulnerabilities and potential therapeutic strategies. Finally, we provide insights on the potential of pan-RAS (ON) inhibition to treat *RAS*- and *KIT*-mutant AM.

## RESULTS

### The collection of acral melanoma samples and models

To contribute to the study of acral melanoma (AM) from Latin American patients, we developed a biorepository of AM samples and models from patients treated at INCA. Seventy-eight tumour samples were collected from surgical resections of AM lesions from 62 patients. A section of sample was used for genomic characterization, while another section was implanted into immunocompromised *NOD/Scid IL2rg^null^* (NSG) mice. Forty of the inoculated samples led to the successful establishment of AM patient-derived xenograft (AM-PDX) models. These were expanded *in vivo* across multiple passages, used to generate cell lines, characterized histopatologically, stored as live tissue, and served as sources of DNA and RNA for molecular analyses (Fig. 1a).

**FIGURE 1:**
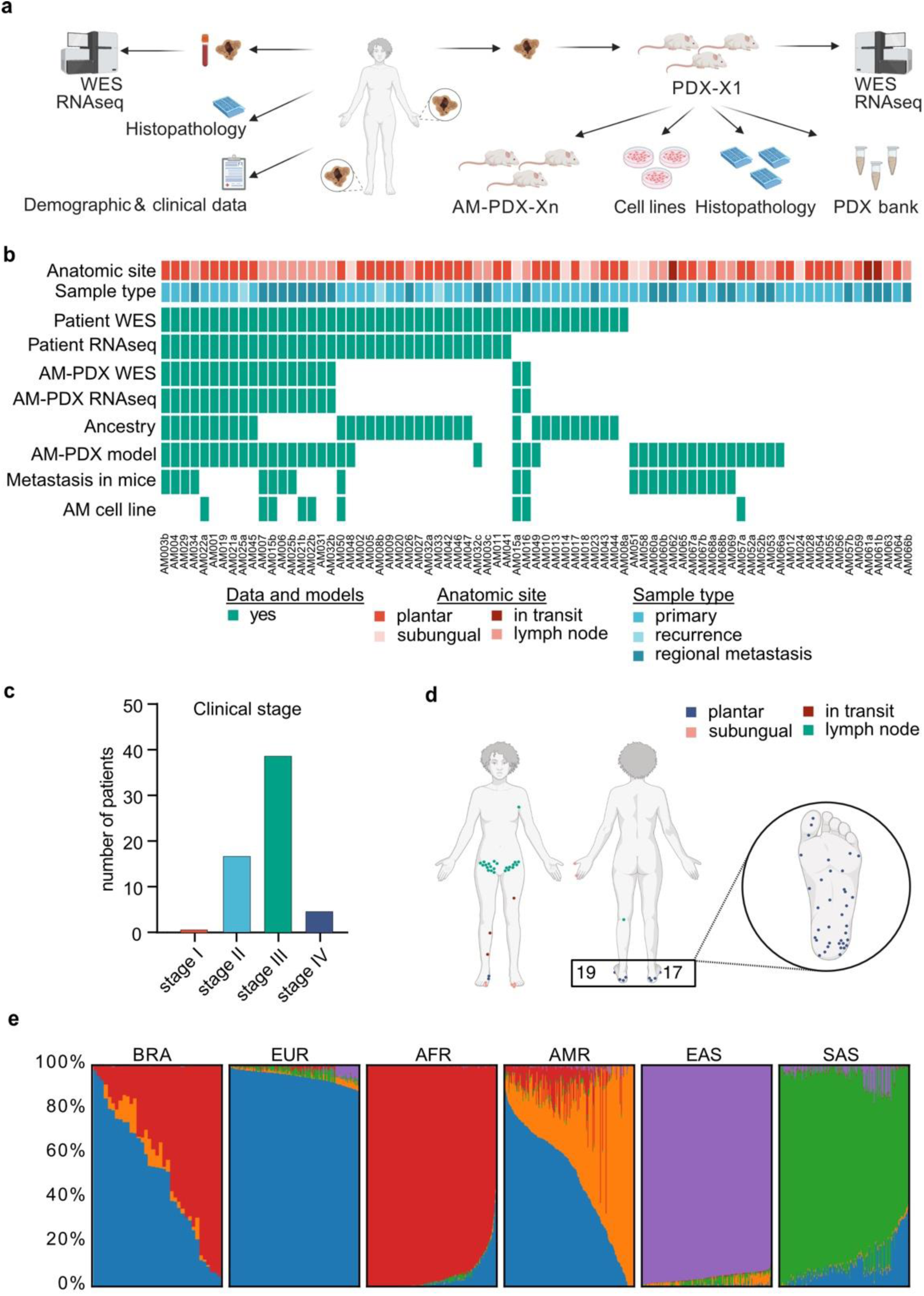
The collection of acral melanoma tumours and models from Brazilian patients. A) Workflow for the generation of a biorepository of AM tumours and patient-derived xenograft (AM-PDX) models. B) Overview of the biorepository showing distribution by clinical characteristics and availability of data and models. C) Bar graphs illustrating patient distribution according to clinical stage (n=62). D) Scheme representing different anatomic locations where samples were collected from. Colours indicate the anatomic site. E) Ancestry estimation for patients included in the study (n=34; BRA). The remaining panels illustrate the superpopulations in the 1000 Genomes Project: European (EUR, blue), African (AFR, red), Amerindian (AMR, orange), and East (EAS, purple) and South Asian (SAS, green). Images in A and D were produced using Biorender.com.

The collection includes primary tumours from both female (58%) and male (42%) donor patients, all of whom consented to participate in this study, 50% of whom were over 70 years old and diagnosed with stage III tumours (63%) at the time of the surgery (Fig. 1b, c, Extended Data Fig.1a, Supplementary Table S1-2). Tumours from plantar and subungual sites, as well as recurrences, in-transit and lymph node metastases from different anatomical locations, were collected (Fig. 1D, Extended Data Fig.1b). The histopathological characteristics of primary tumours, such as the predominance of ulcerated samples and Breslow depth greater than 4mm, are consistent with previously reported characteristics of INCA patients ^28^ and underscore the advanced stage of cases (Extended Data Fig.1c).

To determine whether our cohort reflected Brazil’s ethnic diversity, we analysed genetic ancestry contributions using genotyping data ^37^. The results showed that European and African ancestries accounted for an average of 51% and 44% of the study population’s genetic background, respectively, while Admixed American ancestry contributed approximately 5% (Fig. 1e, Extended Data Fig.1d). Notably, most individuals exhibited an admixed genetic background, primarily a mix of African and European ancestries, consistent with their self-identification (Extended Data Fig.1e) and aligning with Brazilian 2022 census data^38^.

### Genetic profiling of acral melanomas from Brazilian patients

To identify potential driver alterations in AM, whole-exome (WES) and RNA (RNAseq) sequencing data from 40 patients (48 samples) were analysed. Compared with UV-associated CM^8^, the number of mutations observed per sample was low (average: 52.17 across the exome, Supplementary Table S3). Analysis revealed that 27% of patients had mutations in the *NRAS* (15%), *KRAS* (6%) *and HRAS* (6%) genes, 12% in *KIT*, and 6% in *NF1* (Fig. 2a). In our WES cohort, no *BRAF* mutations were observed and mutations in *KRAS/NRAS/HRAS*, *KIT* and *NF1* did not co-occur. The remaining 55% of the tumours, which we named quadruple wild type (4WT), showed no mutations in any of the genes listed above. Additional mutations were found at a lower frequency (9% of the cases) in genes such as *CDKN2A*, *CSMD3* and *LRP1B* (Fig. 2a). We also analysed exon 15 of *BRAF* in the remaining samples, which were not submitted to WES (n=26 samples from 19 patients), by sanger sequencing to determine the frequency of *BRAF*^V600E^ mutation in the complete cohort, identifying V600 mutations in 9.7% of the cases (n=6 patients out of 62 patients) (Extended Data Fig.2a).

**FIGURE 2:**
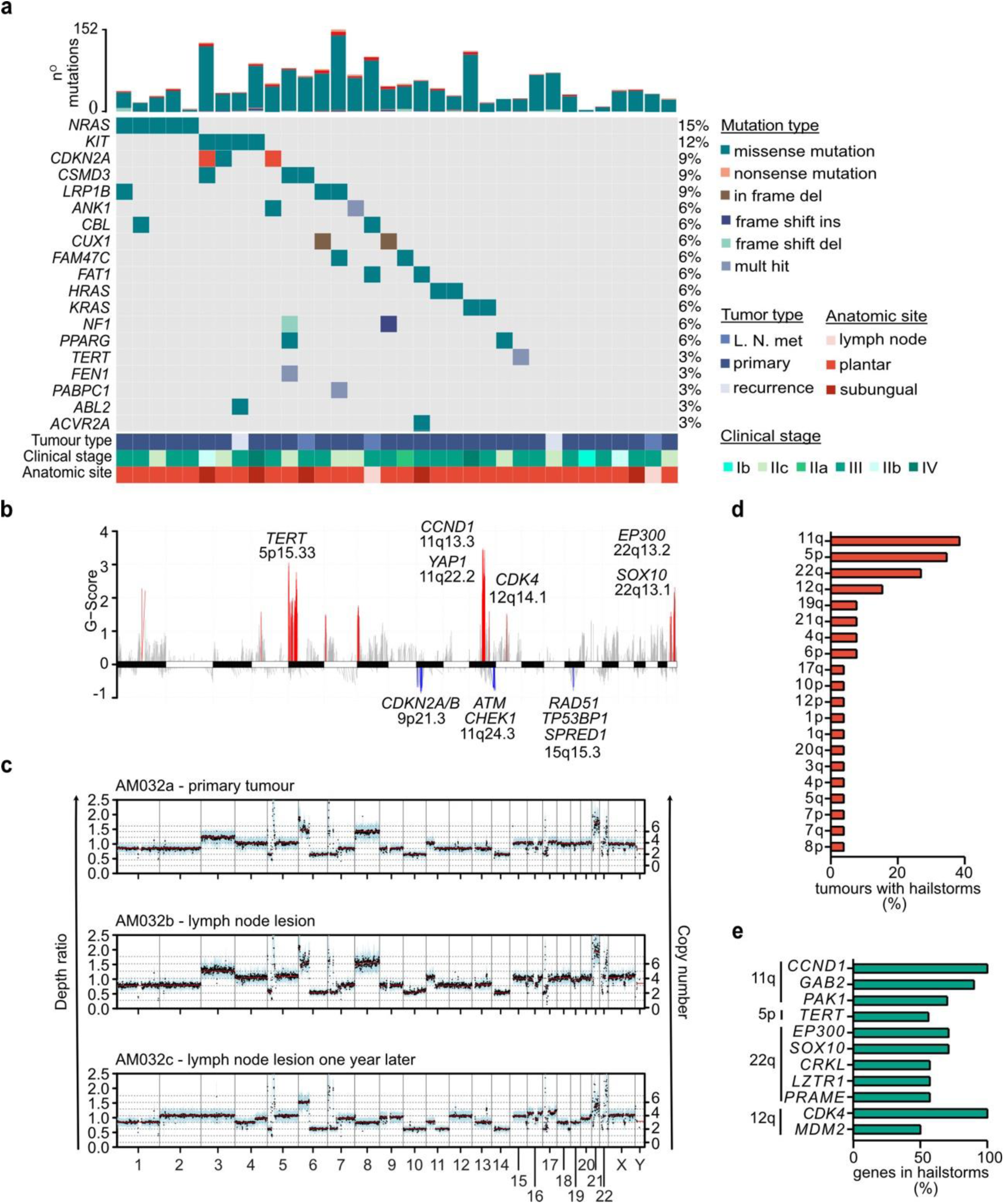
Somatic landscape of acral melanoma in Brazilian patients. A) Oncoplot illustrating the 20 most frequently mutated genes across all samples as identified by CaVEMan and Pindel. Only samples with mutations in these genes are shown (25 samples out of 34, one per patient). The total number of mutations (top) and the frequency of mutations (right) are also shown. B) Distribution of significant regions of amplification (red) and deletions (blue) across chromosomes as identified by Sequenza and Gistic2 in 25 tumours, one per patient. Representative genes are shown along with the respective cytoband. C) Example of tumours with hailstorms. Three samples from patient AM032 are illustrated. The left Y-axis and black dots represent depth ratios (tumour/normal) across different genomic regions. Blue shading represents variability in the depth ratio. The right Y-axis and red lines show the estimated copy number for each chromosome segment. D) Frequency of chromosomal arms affected by hailstorms (n=26 samples). E) Frequency with which specific genes are located within hailstorm boundaries. Only the most frequent hailstorms are illustrated. In panels A, B, and E, only genes catalogued by COSMIC or previously related to melanoma are illustrated.

Copy number alterations (CNA) were highly prevalent. In total, 19 genomic regions spanning different chromosomes were significantly altered across all tumours (q < 0.1; Fig. 2b, Extended Data Fig.2b). Among these, 16 regions harboured amplifications in known melanoma driver genes including *TERT*, *CCND1*, *YAP1*, *CDK4*, *EP300*, and *SOX10*. Significant deletions were less frequent (three regions) and affected tumour suppressor genes involved in key cellular processes such as cell cycle regulation (*CDKN2A/B)*, DNA damage response (*ATM* and *CHEK1*) and DNA repair (*TP53BP1* and *RAD51*). Loss of *SPRED1*, a negative regulator of the MAPK pathway, was also detected. Based on WES-derived estimates, all deletions appeared to be partial (i.e., affecting a single allele).

Most tumours were characterized by the presence of high levels of amplification of certain chromosomal segments, which were generally maintained in other sequenced tumours from the same patient (Fig. 2c). These genomic events, previously termed “hailstorms”, have been identified at high frequency in AM^8^. To further characterize their prevalence, we quantified these events across tumours and found that, consistent with previous studies, they affected 16 chromosomes in a non-random fashion, with 11q events occurring in 38.5% of the tumours, 5p in 34.6%, and 22q in 26.9% (Fig. 2d, Supplementary Table S4). Hailstorms were characterized by the consistent presence of well-established oncogenes (Fig. 2e). The 11q hailstorms consistently harboured *CCND1* (present in 100% of the events), while *GAB2* and *PAK1* were present in 90% and 70% of these events, respectively. Moreover, *TERT* amplification was present in 55% of 5p events, whereas the 22q hailstorms frequently involved *EP300* (86%), *SOX10* (71%), *CRKL* (57%), and *LZTR1* (57%), and 12q hailstorms contained *CDK4* (100%), *MDM2* (75%). Copy number gains showed strong correlation with gene expression levels for most of these genes (Extended Data Fig.3a).

Samples with hailstorms on chromosomal arms 11q and 22q, but not on other arms, including 5p, exhibited a significant increase of focal CNA burden (Extended Data Fig.3b, c) and loss of the key DNA damage response genes *ATM* and *CHEK1* (11q: p=0.03 and 22q: p=0.03; Fisher Exact test), suggesting a potential link between these genomic alterations. Of note, no significant difference in broad and focal CNA burden among the *KRAS/NRAS/HRAS*, *KIT*, *NF1*, or 4WT subtypes was observed (Extended Data Fig.3b, c).

Gene fusions were detected at an average of 4.7 events per tumour, with 2.5 of these preserving the original open reading frame (Extended Data Fig.4a). While every tumour contained at least one in-frame gene fusion, several of which encoding kinases, each fusion was unique to the individual patient (Supplementary Table S5). Several of these fusions involved genes that were significantly amplified across patient tumours, such as *CRKL*::*PI4KA* (AM022a and AM022b), *PAK1::GAB2* (AM033) and *RBM14::YAP1* (AM046), all identified within the hailstorm region of chromosome arm 11q. Notably, most gene fusions, 51.5% when considering both fusion partners and 65.5% when including at least one partner, occurred within hailstorm regions (Extended Data Fig.4b).

Finally, mutational signatures associated with mutations and CNA were also analysed. The dominant pattern was signature 5, a clock-like signature with an unknown aetiology and associated with aging^39^. In half of the samples, there was a minor contribution of signatures 7a and 7b, which are linked to UV exposure^39,40^. Regarding CNA, we observed signatures associated with chromothripsis and chromosomal instability (Extended Data Fig.4c).

### Generation and characterization of the AM-PDX collection

We subcutaneously inoculated NSG mice with tumour fragments to develop AM-PDX models. Out of 78 samples, 40 successfully engrafted, 33 failed, and 5 were interrupted due to adverse events or failure to reach the endpoint criteria of no tumour growth within six months (Fig. 3a). This resulted in an overall success rate of 54.8%. Notably, freshly inoculated samples achieved a success rate of 66.7%, whereas frozen samples reached only 21.0% (Extended Data Fig. 5a).

**FIGURE 3:**
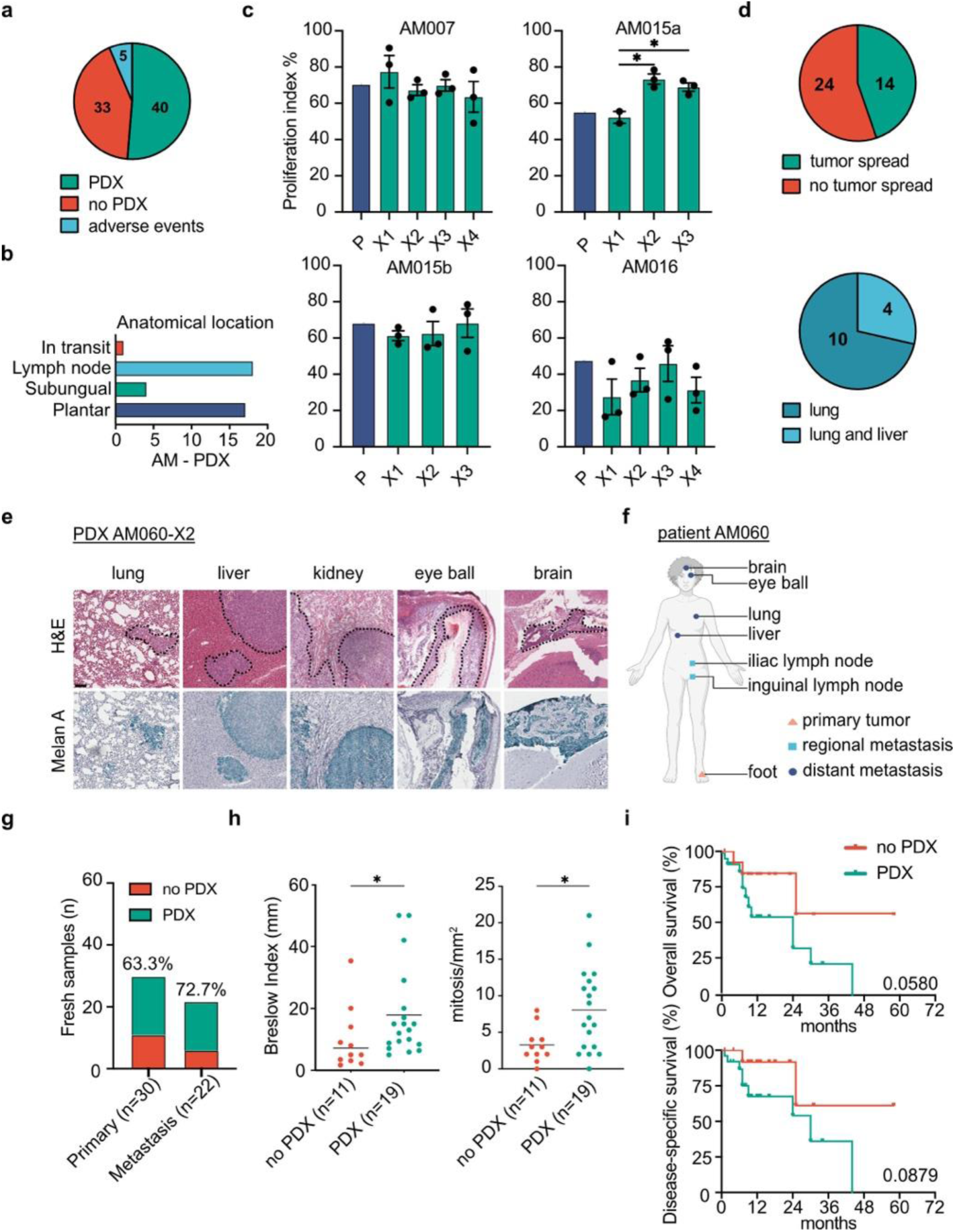
Establishment and characterization of AM-PDX models. A) Proportions of successful generation of AM-PDXs (green), no tumour growth (red), and experiments interrupted due to adverse events (blue) (n=78 implanted tumour samples). B) Anatomical origin of tumours successfully engrafted into mice (n=40). C) Proliferation index evaluated on patient tumours and corresponding AM-PDX models as calculated by the percentage of Ki-67-positive cells in hotspot areas of samples; data shown as mean (+/− SEM). *: p < 0.05; One-way ANOVA with Tukey’s post-test. D) Top pie chart: Proportion of metastases formation in the first passage (X1) of AM-PDX-bearing mice. AM-PDXs with (green), and without metastases (red) are indicated (n=38). Bottom pie chart: Distribution of metastases according to the mouse organ affected (n=14). E) H&E and Melan A staining of sections from different organs of AM060a-X2-bearing mice. Black dashed lines denote tumour areas. Scale bar 100 µm. F) Scheme of primary lesions (orange triangle), regional (light blue square) and distant (dark blue circle) metastases diagnosed in patient AM060 by routine imaging exams. G) Proportions of successful (green) and unsuccessful (red) AM-PDX generation from primary tumour and metastasis samples. Only freshly inoculated samples were considered, (n=52). Success rate is indicated as % on top of the columns. H) Comparison of successful (green) and unsuccessful (red) AM-PDX generation according to sample type (p=0.558, Fisher’s exact test) and histopathological characteristics of primary samples (F: p=0.02, Wilcoxon rank-sum test; G: p=0.013, Unpaired t test, Centre line, median; n=30). I) Kaplan-Meier showing overall survival (OS; p=0.06) and disease-specific survival (DSS; p=0.08) for patients whose tumours successfully engrafted (green, n= 25) compared to those that did not (red, n=14). Log-rank test. For AM-PDX group/No-PDX: Hazard Ratio (HR) of 3.10 (95%CI 1.12 to 8.61) for OS and 3.43 (95%CI 1.04 to 11.30) for DSS.

Half of the models originated from plantar and subungual sites, while the other half were derived from lymph node or in transit metastases (Fig. 3b). *In vivo* growth rates varied among the AM-PDX models, with a median time to reach a palpable size (∼200mm^3^) of 11 weeks considering all inoculated samples (Extended Data Fig.5b). Histopathologically, AM-PDX models closely resembled the original patient tumours (Extended Data Fig. 5c), with no differences in proliferation rates as measured by Ki-67 positivity (Extended Data Fig. 5d). Serial passaging in mice preserved the tumours’ histopathological structure, including cell morphology, Ki-67 expression, and mouse cell infiltration (Fig. 3c, Extended Data Fig.5e, Extended Data Fig. 6).

To evaluate the metastatic potential of AM-PDX models, we collected the lungs and livers of AM-PDX-bearing mice at the end of the experimental endpoint. These were analysed for the presence of human tumour cells by H&E staining and immunohistochemistry on one representative tissue section of each organ. Metastatic spread was detected in 14 out of 38 (36.8%) first passage (X1) AM-PDX models analysed, all of which affected the lungs (Fig. 3d). In addition, four of the models with metastatic spread (28.5%) showed metastatic foci in the mouse liver. Analysis of additional passages of the AM-PDX models revealed dissemination to other organs, with one case showing metastases to the kidneys, brain, and eyes in addition to lungs and liver (Fig. 3e). Interestingly, the patient from whom this tumour was derived had developed distant metastases to the eyes as well, with the kidney being the only site affected uniquely in the AM-PDX model (Fig. 3f).

To determine whether more aggressive tumours were more likely to establish AM-PDX models, we analysed the association of clinical or histopathological characteristics of patient tumour samples with tumour take. To eliminate any bias caused by loss of viability due to sample freezing, only freshly inoculated samples were included in the analysis. There was no difference in tumour take between primary tumour and metastasis samples (Fig. 3g). However, among the primary tumours, patient samples that successfully engrafted had greater Breslow depth and higher mitotic index compared to those that did not engraft (Fig. 3h) (Wilcoxon rank-sum test, p=0.02; Unpaired t test, p=0.01, respectively). When evaluating survival outcomes, we observed that the median overall survival (OS) and disease-specific survival (DSS) for the group whose tumour samples successfully engrafted were 24 and 30 months, respectively. In contrast, for the group whose tumour samples failed to engraft, the median was not reached at the end of the follow-up period (58 months) (Fig. 3i). However, these differences were not statistically significant (log-rank test, OS: p=0.06; DSS: p=0.09). Together, these findings indicate that more aggressive tumours are more likely to successfully establish an AM-PDX.

### AM-PDX models maintain the genetic structure of the corresponding patient tumours

To validate the fidelity of the AM-PDX models to the original patient tumours, we analysed the exomes and mRNA of the first passage AM-PDX models (X1) and compared them to the corresponding patient tumours. Results showed a strong correlation between AM-PDX models and their matched patient tumours in both mutation burden and CNA (Fig. 4a). On average, 73% of the mutations and 67% of the in-frame gene fusions detected in the patient tumours were retained in the AM-PDX models, although some events were either newly acquired or lost (Fig. 4b, c). The overall pattern of CNAA across all chromosomes was largely consistent with the corresponding patient tumour (Fig. 4d). Similarly, the presence and distribution of hailstorms across chromosomes remained consistent (Fig. 4d, Extended Data Fig.7). As a result of the preserved genetic structure of the models, driver genes frequently altered by mutations or CNA were replicated in the models (Extended Data Fig.8a).

**FIGURE 4:**
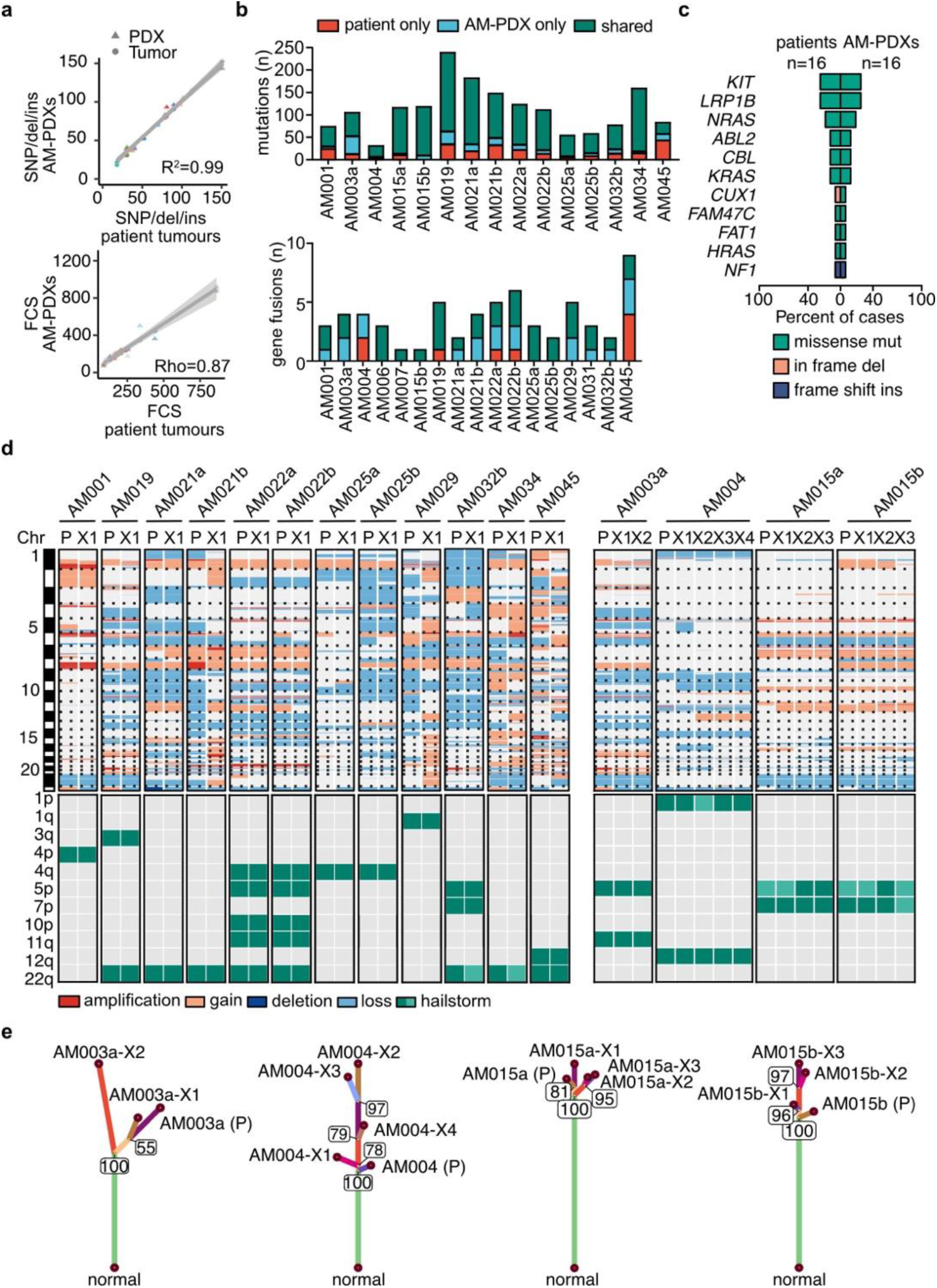
AM-PDXs recapitulate the genomic structure of the corresponding patient tumours. A) Pearson correlation of SNVs, deletions and insertions between AM-PDXs and patient tumours (r= 0.99, IC95%: 0.97-0.997, p=1.96×10^−13^). Spearman correlation of the focal copy number scores (FCS) between AM-PDXs and patient tumours (rho=0.87, S=58.5, p=4×10^−5^). Scores were determined using the CNApp tool and the grey line represents the best fit line with 95% CI. B) Number of mutations and gene fusions found only in patient tumours (red), only in AM-PDXs (blue) or in both (green). C) Frequency of the 20 most frequently mutated genes from COSMIC gene census identified in AM-PDXs and the corresponding patient tumours. Mutation type is shown (n=16 of each, patient sample vs PDX-X1 sample). D) Heatmaps illustrating the copy number variation across all chromosomes of the patient tumour samples (P) and their corresponding AM-PDX models (Xn). On the top, colours indicate the type of alteration as identified by Sequenza and Meskit and chromosomes are separated by horizontal dotted lines (n=16; P vs Xn pairs). At the bottom, hailstorms identified are illustrated according to chromosome cytoband affected. Light green boxes indicate hailstorms that were detected by visual inspection after event calling (n=16 of each, patient sample vs PDX-X1 sample). E) Phylogenetic trees illustrate changes in somatic SNV and indels in AM-PDXs and patient tumours as identified by Sequenza and Meskit. Four models are shown. Branch length is proportional to somatic mutations detected in samples.

Phylogenetic analysis of sequential *in vivo* passaging in representative AM-PDX models revealed limited emergence of new detectable mutations, which may reflect clonal selection over time. However, the total number of acquired mutations was low, on average below 100 events (Fig. 4e, Extended Data Fig.9, Supplementary Table S6). Importantly, main driver alterations were retained throughout passaging (Extended Data Fig.8b, Supplementary Table S7). Together, these findings demonstrate that AM-PDX models faithfully replicate the key histological and genetic characteristics of patient tumours across *in vivo* passaging. They provide a robust platform for studying AM biology and uncovering tumour vulnerabilities.

### Patterns of drug sensitivity in acral melanoma cells

Building on the robust genetic and histopathological fidelity of the AM-PDX models to their corresponding patient tumours, we utilized these models to investigate therapeutic vulnerabilities in AM. We screened AM-PDX-derived cell lines with a panel of potential pharmacological anti-cancer agents that have been extensively studied in preclinical models and clinical trials (Fig. 5a). In addition to inhibitors of established melanoma drivers frequently altered in AM, such as CDK4, MDM2, KIT and MAPK, we included agents that interfere with cell cycle regulation, DNA damage response and repair (DDR), protein stability and epigenetic regulation. This rationale was based on the hypothesis that the high burden of genomic alterations could render these cells sensitive to inhibition of key regulators of these cellular processes.

**FIGURE 5:**
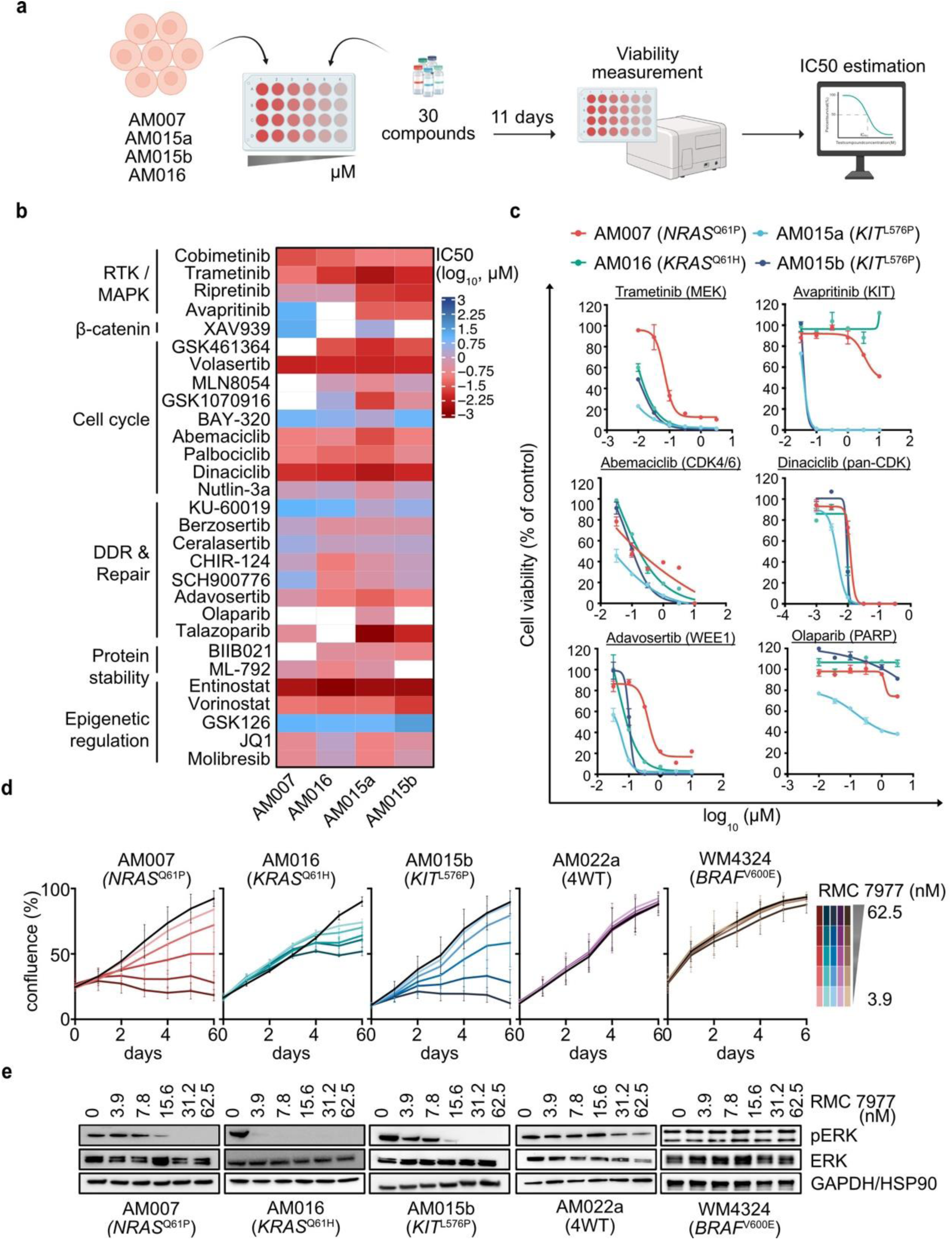
Patterns of drug sensitivity in acral melanoma cells. A) Overview of the pharmacological screens performed in AM-PDX-derived cell lines. Scheme was produced using Biorender.com. B) Cells were treated with 30 small-molecule inhibitors for 11 days at different concentrations. Cell viability was measured using ATPlite and IC50s were calculated. Heatmap shows the log_10_-transformed IC50s with drugs grouped according to their functional classification. White indicates cases where a drug failed to achieve 50% effectiveness (cell viability loss) in each cell line. C) Representative dose response curves of inhibitors included in the screen. D) Cells were treated with DMSO (black line) or increasing concentrations of RMC-7977 and monitored using the Incucyte live cell imager for 6 days. Pictures were taken every 24 hrs and treatment was renewed after 3 days. Mean values of 3 independent biological replicates were plotted. E) Western blots of cells treated with the indicated doses of RMC-7977 or DMSO for 2 hrs. HSP90 (WM4324) and GAPDH (AM007, AM016, AM015b, AM022a) were used as loading controls. A representative of 3 independent experiments is shown.

Four cell lines were screened with different doses of 30 inhibitors (Supplementary Tables S8 and S9). Most drugs demonstrated nano- or micromolar IC50s for all cell lines (Fig. 5b, Supplementary Table S7), although sensitivities varied. For instance, MEK and inhibitors were consistently effective (IC50 < 0.1 µM) while responses to KIT inhibitors ripretinib and avrapritinib were markedly effective in *KIT*^L576P^-mutant cell lines AM015a and AM015b (IC50 < 0.05 µM), but not in *NRAS^Q61P^* (AM007) and *KRAS^Q61H^*(AM016) - mutant cells (IC50 not reached). Among DDR inhibitors, adavosertib was the most effective (WEE1 inhibitor; IC50 < 0.5 µM), whereas olaparib (PARP inhibitor) had limited effect and failed to reach an IC50 in three out of the four cell lines tested (Fig. 5c, Extended Data Fig.10). These results reveal vulnerabilities that can be further explored in AM and underscore the need for personalized therapeutic approaches.

Recently, the pan-RAS (ON) inhibitor RMC-7977 has demonstrated efficacy in preclinical models of non-small cell lung cancer, colorectal cancer, and pancreatic adenocarcinoma with *KRAS/NRAS/HRAS* mutations, both *in vitro* and *in vivo*^41^. To test the potential of RMC-7977 in AM, we treated AM cell line harbouring *NRAS^Q61P^* (AM007), *KRAS^Q61H^* (AM016), and *KIT*^L576P^ (AM015b) mutations, in addition to a 4WT AM cell line (AM022a) and a *BRAF*^V600E^-mutant CM cell line (WM4324). The results revealed that RMC-7977 effectively decreased the viability of *NRAS^Q61P^*, *KRAS^Q61H^*, and *KIT*^L576^–mutant cells (Fig. 5d, S11a, b), at least partially via increased cell death (Extended Data Fig.11c). These effects were accompanied by MAPK pathway suppression in sensitive lines, while 4WT and BRAF^V600E^-mutant cells were unresponsive and had little effect in blocking the MAPK pathway (Fig. 5e). These findings highlight the therapeutic potential of RAS inhibition in AM.

### CRISPR/Cas9 Knockout screens identify acral melanoma-specific dependencies

To identify additional vulnerabilities of *NRAS^Q61P^* and *KRAS*^Q61H^ -mutant AM cells, we performed CRISPR knockout screens (Fig. 6a). For that, AM007 and AM016 cells expressing active spCas9 were transduced with a genome-wide gRNA library^42^ and analysed for cell fitness at two time points. Bayesian Analysis of Gene Essentiality^43^ scored *NRAS* and *KRAS* as true hits in their respective mutant contexts, serving as positive controls for the screens (Fig. 6b). Next, we examined shared vulnerabilities, identifying 822 genes whose loss decreased cell viability in both cell lines (Extended Data Fig.S11d, Supplementary Table S10). Of these, 461 genes were classified as known common dependencies, which are essential for cell viability in nearly all human cell lines and are mostly involved in fundamental cellular processes^44^. Indeed, among those genes were several that encode kinases involved in cell cycle regulation and DDR, including *PLK1*, *AURKB*, *ATR*, *CHK1*, and *WEE1* (Extended Data Fig.S11e), which were also identified as potential vulnerabilities in the drug screen.

**FIGURE 6:**
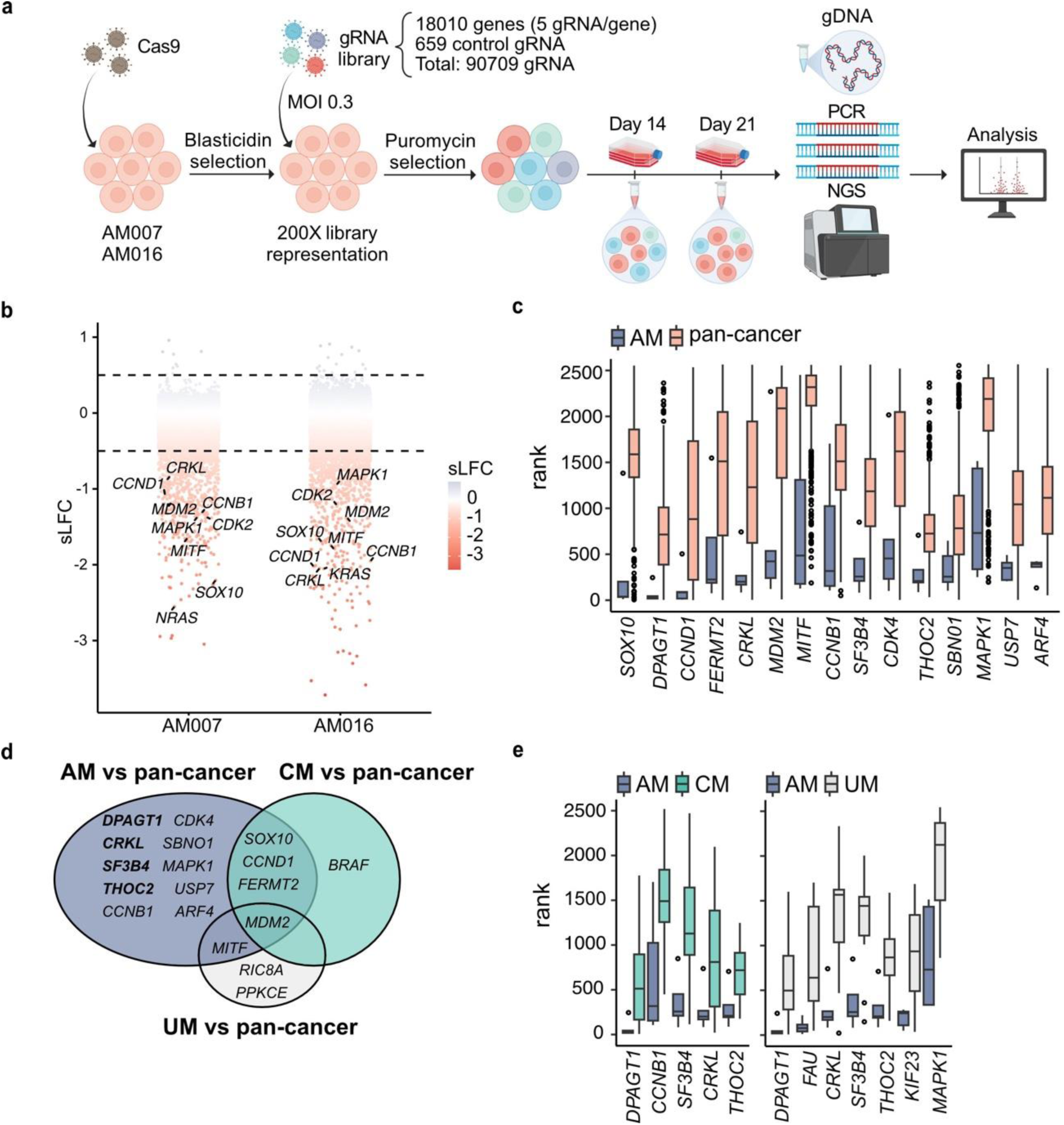
CRISPR/Cas9 knockout screens identify AM vulnerabilities. A) Overview of the genome-wide CRISPR knockout screens performed in AM-PDX-derived cell lines. Scheme was produced using Biorender.com. B) Waterfall plot illustrating genes identified as common dependencies in AM007 and AM016 through BAGEL analysis, ranked by their scaled Log Fold Change (sLFC). Selected genes are shown. Dotted lines indicate a significant threshold (0.05 < sLFC < −0.05). C) Comparison of genes identified as essential in acral melanoma (AM, n=5 cell lines) and other tumour types (pan-cancer, n=1010 cell lines) using DepMap datasets (p < 0.05, Mann-Whitney U test). D) Venn diagram showing the overlap of specific dependencies identified in acral, cutaneous, and uveal melanoma, as described in panels C and D. E) Comparison between genes identified as essential in AM (n=5 cell lines) with genes in cutaneous (CM, n=56 cell lines) and uveal melanoma (UM, n=10 cell lines) using DepMap datasets (p < 0.05, Mann-Whitney U test). For panels C and D, only genes identified as more essential in AM are shown. Box plot features: centre line = median; box limits = upper and lower quartiles; whiskers = 1.5x interquartile range; points = outliers.

Among the remaining 361 genes were key regulators of melanoma biology, including *MITF* and *SOX10*, and *CRKL*, which is frequently amplified in AM and implicated in anatomical distribution and growth of AM cells^10,45^ (Fig. 6b). *MAPK1*, encoding ERK2, a central enzyme in the MAP kinase pathway, also emerged as a significant dependency. Additionally, several cell cycle-related genes, including *CCNB1* and *CCND1*, which encode Cyclin B1 and D1, respectively, together with *MDM2* and *CDK2*, were identified.

To explore AM gene essentiality more comprehensively, we leveraged genome-wide CRISPR knockout screen data from the DepMap portal, which included data from three additional AM cell lines (WM3211, M040416 and MM160113), and integrated it with AM007 and AM016 data (n=5 AM cell lines). Comparison of AM cells with cells from multiple non-melanoma cancer types (pan-cancer) revealed 15 genes with increased essentiality in AM. *MITF*, *SOX10*, *MDM2*, *CDK4* and *CRKL* were among the significant hits (Fig. 6c). However, none of these genes passed the p value correction for multiple comparisons. Although this is likely due to the low number of AM cell lines available, we cannot rule out that some of these hits were false positives.

To mitigate this limitation, we compared the gene essentiality scores of non-acral cutaneous melanoma (CM) and uveal melanoma (UM) cells to those of the same pan-cancer cell line panel. This revealed that CM cells were significantly more dependent on *SOX10* and *BRAF*, along with *CCND1*, *MDM2*, and *FERMT2*, than cells derived from other cancer types even after p value correction. *MDM2*, *RIC8A, PRKCE*, and *MITF* were identified as more essential in UM cells compared to pan-cancer (Extended Data Fig.11f). By overlapping the dependency lists from the pan-cancer comparisons, we uncovered both shared and subtype-specific dependencies (Fig. 6d). Interestingly, four out of 15 genes identified as more essential in AM compared to non-melanoma cancer cells were also found to be more essential in AM compared to CM or UM (Fig. 6d). In addition to *CRKL*, which has a well-known role in AM biology^45^, *SF3B4*, *THOC2*, and *DPAGT1* were identified as AM essentialities.

Based on these findings, we hypothesized that expression of *CRKL*, *SF3B4*, *THOC2*, and *DPAGT1* might distinguish AM cells and reflect subtype-specific vulnerabilities. To assess this, we integrated publicly available single-cell RNA-seq (scRNAseq) datasets consisting of six AM and three CM primary tumours ^46^. We observed that expression levels of all four genes were higher in melanoma cells compared to other cell types present in the tumour microenvironment (Fig. 7a, b). In melanoma cells alone (Fig. 7c), differential expression analyses revealed that *SF3B4* levels were higher in AM than CM, whereas no difference was seen for *CRKL*, *THOC2*, and *DPAGT1* (Fig. 7d).

**Figure 7:**
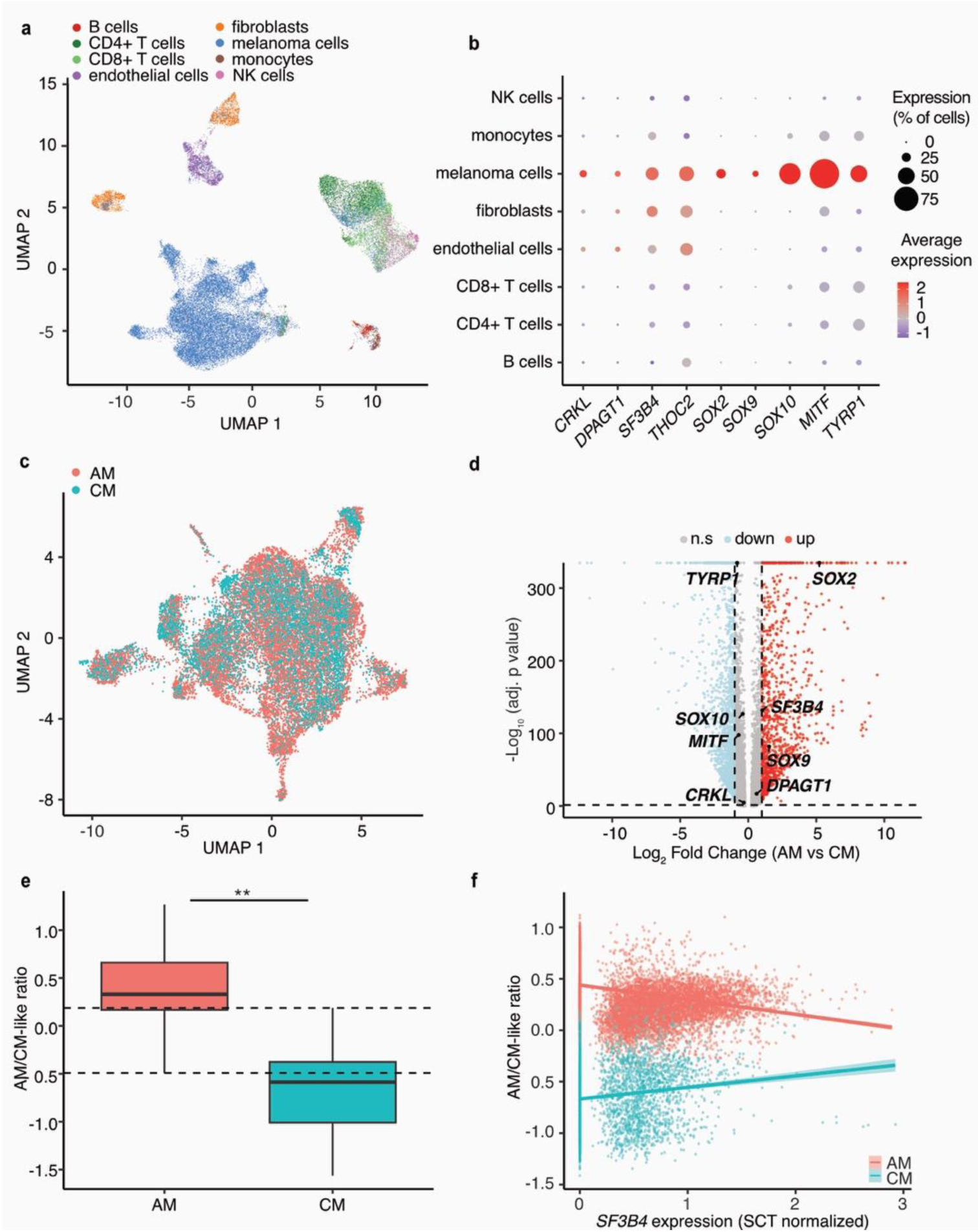
*SF3B4* expression correlates with melanoma-defining transcriptional profile. A) UMAP visualization of scRNA-seq data showing cell distribution of primary acral (AM, n=6) and cutaneous (CM, n=3) melanoma samples, coloured by the annotated cell types. B) Gene expression levels across cell types. Dot size represents the percentage of cells expressing the gene, and colour indicates the average scaled expression value. C) UMAP plot of melanoma cell cluster after the integration of AM and CM samples using Harmony coloured by dataset. D) Volcano plot of differential gene expression in AM vs CM in melanoma cells using the MAST test. Genes coloured in red are significantly up-regulated in AM, and those coloured in blue are down-regulated. Not significant genes (n.s.) were coloured in grey. E) The ratio of the average expression of AM-like and CM-like signatures was calculated using the top 100 specifically differentially expressed genes of the melanoma cluster (median= 0.34 and −0.62, respectively; Two-sided Wilcoxon rank-sum test, p < 2.2 × 10^−16^). The dotted line indicates the overlap ratio regions. F) Linear model analysis of the relationship between *SF3B4* expression and AM/CM-like ratio in AM (red) and CM cells (blue). The interaction between *SF3B4* expression and melanoma subtype was evaluated (p < 2 × 10^−16^; F (3,19) = 1.78 × 10^4^, p < 2.2 × 10^−16^, Adjusted R^2^ = 0.734).

Since previous work has shown that melanocytes residing in acral skin are phenotypically different from those present in sun-exposed body areas^47^ we next generated AM-like and CM-like gene expression signatures composed of the top 100 differentially expressed genes within the scRNAseq datasets (Fig. 7e; Supplementary Table S11). Notably, *SF3B4* was captured within the AM-like signature with expression levels significantly higher in these cells (MAST test, padj < 2.2e-16; average log_2_FC = 1.0525) when compared to CM-like cells, a result further confirmed by linear modelling (Fig. 7f; t test, p < 2 ×10^−12^). These results suggest a distinct role for *SF3B4* in AM associated with enriched expression and gene essentiality.

## DISCUSSION

In this study, we provide one of the most comprehensive analyses of AM tumours and models to date, particularly from a Latin American population with substantial ancestral admixture. Our biorepository of patient-derived xenografts (AM-PDX) and cell lines, as well as biospecimens of AM from Brazilian patients, contributes to global efforts to characterize AM in diverse populations. This provides a unique resource for studying the molecular mechanisms driving disease progression and potential actionable therapeutic vulnerabilities of a subtype of melanoma that is both clinically aggressive and significantly understudied.

Genomic analysis of tumours revealed hallmark features of AM, including relatively low mutation burden and CNA burden. Consistently, tumours exhibited minor contributions from UV-related signatures and several signatures suggestive of drastic genomic structural events, such as chromothripsis and hailstorms. In contrast to UV-related melanoma, where *BRAF*^V600E^ alterations dominate, more than half of tumours lacked mutations in either *BRAF* or other canonical MAPK driver genes such as *KRAS*, *NRAS*, *HRAS*, *KIT*, and *NF1*. In addition, the frequency of *BRAF* mutations observed in our cohort (9.7%) was notably lower than studies from AM cohorts in Australia and the UK, which can be close to 20%^7,10,48–50^. Instead, the tumours showed widespread chromosomal amplifications, often in patterns known as hailstorms, clustered high amplitude copy number gains^8^, which were most frequent on chromosome arms 11q, 5p, and 22q. These hailstorms consistently involved oncogenes such as *CCND1*, *TERT*, and *CDK4*, whose amplifications were strongly correlated with elevated gene expression, suggesting they play a central role in AM pathogenesis. Our findings closely parallel observations from recent studies in Asian, Mexican and multi-ethnic cohorts^7,8,10,12,13,50^ reinforcing the concept that AM relies more on structural variations than on point mutations for malignant progression.

Interestingly, we found that more than half of the gene fusions occurred within hailstorm regions, often involving amplified oncogenes. These findings suggest a dynamic interplay between amplifications and rearrangement events in AM and further studies involving larger cohorts and integrative genomic analyses are needed to elucidate the underlying mechanisms driving these complex events.

The observed association between hailstorms on chromosomal arms 11q and 22q, the recurrent loss of *ATM* and *CHEK1*, and the elevated burden of focal CNA collectively suggest a deficiency in genomic maintenance in AM. Loss of *ATM* and *CHEK1*, central regulators of DDR signalling, can impair cellular response to double-strand breaks and replication stress, fostering genomic instability and facilitating the emergence of highly focal, clustered amplifications characteristic of hailstorms^51–53^. In AM, recent genomic studies have highlighted the prominence of complex rearrangements and focal amplifications, including hailstorms and tyfonas, amplification-driven rearrangements involving extrachromosomal-like structures or breakage-fusion-bridge cycles, as core tumorigenic mechanisms^8,54,55^. These findings suggest that hailstorms may not be random structural artifacts but rather the consequence of a permissive genome structure, offering both mechanistic insight and potential therapeutic opportunities, particularly for agents targeting replication stress or synthetic lethal pathways.

The AM-PDX models that we established faithfully preserved both the histopathological and genetic hallmarks of their respective tumours across passaging, including driver mutations, gene fusions and CNA patterns. In particular, hailstorm patterns present in the patients’ primary tumours were reproduced without attenuation in the corresponding AM-PDXs. Coupled with our observation that the same genomic architecture persists in matched lymph-node metastases, these data suggest that such complex structural events arise early in AM evolution and remain stable during progression. This conclusion is consistent with recent evidence showing that hailstorms are already detectable in *in situ* AM lesions and typically endure throughout the disease course^8^.

Using cell lines derived from these AM-PDX models, we performed genetic and pharmacological screens. The CRISPR/Cas9 knockout screens uncovered a set of shared and AM-selective gene dependencies, many of which overlapped with the drug screen results. Core melanoma transcription factors such as *MITF* and *SOX10* were essential, consistent with their roles in melanocyte differentiation and survival^8^. Notably, genes involved in cell cycle regulation (*CCND1*, *CCNB1*, *CDK2*) and p53 regulation (*MDM2*) also emerged as both essential in the CRISPR screens and as effective drug targets. Inhibitors targeting CDK4/6 and MDM2 demonstrated consistent and potent cytotoxicity across AM-PDX-derived cell lines, underscoring the functional relevance of these genetic dependencies. Likewise, *WEE1*, which showed strong dependency signals in the CRISPR screens, was pharmacologically validated through sensitivity to the WEE1 inhibitor adavosertib.

*CRKL*, frequently amplified in our cohort and identified as an AM-specific dependence, represents a particularly compelling therapeutic candidate. CRKL has been implicated in tumour growth and invasion in other cancers and in AM, it was shown to play a role in the anatomic location of the tumours^45,50^. Here we show that its essentiality in AM represents a novel finding with potential therapeutic relevance. While direct CRKL inhibitors are not yet available, its role as an adaptor in the RAS-MAPK signalling axis suggests it may modulate sensitivity to MAPK-targeted agents. In line with this, RAS pathway inhibition using the pan-RAS (ON) inhibitor RMC-7977 was effective in *NRAS*, *KRAS*, and even *KIT* -mutant AM cell lines, which also showed MAPK pathway dependence in the CRISPR screen data.

This concordance between drug-induced vulnerabilities and genetic dependencies not only reinforces the robustness of our findings but also highlights potential avenues for therapeutic intervention. Importantly, several of these targets (e.g., KIT, MEK1, CDK4, MDM2, WEE1) are already druggable with compounds being tested in clinical trials or approved for other indications, facilitating the translational potential of these insights for AM patients. Further exploration of these compounds alone and in combination with other therapeutic agents in a broader AM cell line panel representative of all AM genomic subtypes is warranted.

Among the more novel and dependencies with greater relevance for AM identified in our CRISPR screens were *SF3B4, THOC2* and *DPAGT1*. The selective dependency on these genes in AM cell lines suggests they may represent previously unrecognized therapeutic vulnerabilities, and further exploration of this potential is needed.

SF3B4 is a core component of the spliceosome and has been associated with aberrant splicing and oncogenesis in hepatocellular carcinoma and pancreatic cancer^56,57^. Interestingly, loss of *SF3B4* affects the development and migration of neural crest cells^58^, which is melanocyte’s cell of origin. Moreover, studies in *Xenopus laevis* models have shown that *SOX10* expression is downregulated following *SF3B4* loss, particularly at the neural plate border, where neural crest progenitors originate^59^. To date, there is no evidence that melanocytes from acral skin follow a distinct differentiation trajectory or undergo branching from those in sun-exposed body sites. However, our findings raise the intriguing possibility that SF3B4 may influence acral melanocyte cell fate, as well as the development and progression of AM. Given its known role in RNA splicing, SF3B4 may regulate the activity of genes critical to acral melanocyte biology and tumorigenesis. Uncovering such regulatory mechanisms may be key to understanding the fundamental biological differences between AM and CM.

In conclusion, through integrated genetic profiling, CRISPR knockout screens and drug screens, we characterized the genetic profile of AM in an admixed Brazilian population and demonstrated that experimental models derived from these patients faithfully recapitulate the tumour characteristics and capture the vulnerabilities of AM. This platform can help uncover genes specifically relevant for AM cell fitness, providing the foundation to study AM biology, to develop effective therapeutic targets, and ultimately offering new avenues for addressing the unmet clinical needs of patients with AM. The identification of *SF3B4* as an AM dependence further attests the utility of these models. This AM-PDX platform can be further used to explore drug combinations and investigate mechanisms of drug resistance. Additionally, the metastatic dissemination patterns observed in our AM-PDX models offer opportunities to dissect the mechanisms of metastatic dissemination and organotropism. Expanding the repertoire of AM models to cover additional genomic subtypes and ancestries will further enhance their utility and inform the design of tailored therapeutic strategies for this aggressive melanoma subtype.

## METHODS

### Ethics Approvals and patient recruitment

This study was approved by the Institutional (CEP) and National (CONEP) Research Ethics Committees (CAAE 98752818.6.0000.5274) including patient recruitment and informed consent, the generation of patient-derived xenografts biorepository, collection of demographic, clinical and histopathological data and molecular and functional analyses. All animal procedures, including the generation of AM-PDX models, were approved by the Institutional Animal Experimentation Ethics Committee (Protocol 003/18). Patients were recruited during their first visit to Hospital do Câncer II (INCA) by a specialized nurse from the INCA Tumour Bank. After receiving detailed information about the study, procedures, risks and benefits, those who agreed to participate signed an informed consent form and completed a socio-demographic questionnaire. Only patients aged 18 or older at the time of recruitment were eligible. Tumour samples were collected from surgical resection of primary tumours, regional metastases, and/or recurrences between March 2019 and May 2022. Blood samples were collected during routine hospital visits for standard blood tests. Clinical data were extracted from medical records and reviewed by the medical staff. Histopathological tumour characteristics were reviewed by a pathologist following the 8^th^ edition of the American Joint Committee on Cancer (AJCC) guidelines^60^. All samples were processed following strict confidentiality protocols in compliance with ethical guidelines for anonymized data sharing. The sample IDs use throughout the manuscript are anonymized identifiers used exclusively within this study and cannot be link to individual patient identities.

### Sample processing

Samples were processed on the same day of the surgery. Part of the surgical sample was collected in a transport medium, composed of RPMI 1640 supplemented with 1% Amphotericin B and 2% Penicillin-Streptomycin 10,000 U/mL and brought to the laboratory for xenografting into mice. The other part was frozen in liquid nitrogen and stored at −20 °C for subsequent DNA/RNA extraction (Allprep DNA/RNA extraction mini kit, QIAGEN). Blood samples were stored at −20 °C for DNA extraction (DNAeasy blood and tissue kit, QIAGEN). All protocols were performed at INCA according to the manufacturer’s instructions.

### Generation of AM-PDX models

Surgical samples stored in transport medium were submitted to a short decontamination protocol including wash steps in 70% ethanol, HBSS containing 1% Amphotericin B and 2% Penicillin-Streptomycin and RPMI 1640 supplemented with 10% FBS, 2% Amphotericin B, and 4% Penicillin-Streptomycin. After processing, tissue fragments were manually cut into 1-3 mm² fragments and immediately grafted in mice or cryopreserved in FBS 10% DMSO.

Male or female NOD/SCID/IL-2Rγ^null^ (NSG) mice, aged 5-10 weeks, were anesthetized with isoflurane at a concentration of 4% vol/L oxygen using an anaesthetic induction vaporizer (XGI-8 Gas Anaesthesia System, PerkinElmer). After shaving, their backs were aseptically prepared with 70% alcohol and 3 to 5 tissue fragments were dipped briefly into Matrigel® Growth Factor Reduced (Corning) and implanted subcutaneously into the flank of each animal (3–5 animals per sample) using an 11G x 3-1/4” cannula (Cadence Science). Tumour growth was monitored weekly by calliper measurement, with tumour volume calculated as follows: the shortest diameter^2^ x the largest diameter / 2.

Tumour onset was defined as the duration from implantation to the development of a palpable tumour (∼ 200 mm^3^). When the subcutaneous tumour reached the experimental endpoint (350-500 mm³), mice were euthanized in a CO₂ chamber at 4% concentration until the complete absence of respiratory movements and motor reflexes was confirmed. Tumours, lungs, liver, and organs with macroscopic suspicion of metastases were collected. Tumours were cut into fragments and snap frozen for molecular analysis, prepared for histological analysis, cryopreserved in FBS 10% DMSO, or inoculated in mice for *in vivo* passaging following similar procedures.

AM-PDX generation was considered successful if at least one animal developed a tumour volume > 350mm^3^; failure was defined as no tumour grow within six months in at least three animals. Experiments were interrupted in cases of adverse events, including unexplained death or signs of animal discomfort such as > 20% weight loss, pain, dehydration, impaired mobility or feeding, and prone positioning.

### Generation of cell lines

AM cell lines were generated from AM-PDXs through mechanical and enzymatic dissociation using 50u/mL DNase I, 200u/mL collagenase IV, and 5mM CaCl_2_ and initially cultured in Mel2% medium consisting of a 1:4 mixture of MCDB153 to L15 supplemented with 2% FBS, 5 μg/ml Bovine Insulin, 15 μg/ml BPE, 5 ng/ml EGF and 1.68 mM CaCL_2_. After stable growth was achieved (over 10 passages), cells were transitioned to RPMI 1640 supplemented with 10% FBS, 2 mM L-glutamine, 1 mM sodium pyruvate, and 10000 U/mL penicillin-streptomycin to simplify culture conditions. Cells were cultured at 37 °C with 5% CO₂ and tested regularly to ensure they were free of mycoplasma contamination (MycoAlert® PLUS Mycoplasma Detection Kit, Lonza).

### Immunohistochemistry analyses

Patient and AM-PDX samples were fixed in buffered 10% formalin and embedded in paraffin blocks. Sections (3 μm) were prepared for haematoxylin and eosin (H&E) staining or immunohistochemistry (IHC) analysis using Trilogy buffer (Cell Marque) in a pressure cooker to perform deparaffinization, dehydration and heat-induced epitope retrieval. Novolink Polymer Detection Systems kit (Novocastra, Newcastle-upon-Tyne) was used following the manufacturer’s instructions. Anti-MelanA (clone EP1422Y, Abcam, ab51061, 1:1000 dilution) and HMB45 (Roche, 90-4366) antibodies were used as melanoma markers. Anti-Lamin A/C (clone: BEO-12, Boster Bio, M00438, 1:12000) was used as a human-specific cell marker. Proliferation index was evaluated using anti-ki-67 (clone MIB-1, Dako, 1:500 dilution). The visualization of antigen-antibody binding sites was produced using a PolyDetector HRP Green Substrate-Chromogen Kit (Bio SB). Histology slides were scanned using the Aperio CS2 Digital Slide Scanner (Leica Biosystems) and images were imported into Slide Viewer software (3D Histech).

Patient and AM-PDX samples were analysed for cell morphology, melanocytic origin and proliferation index. For cell morphology, ten fields of the H&E-stained slides were selected, and tumours were classified as spindle when >50% of each field analysed, epithelioid when >50% of each field analysed, and mixed when similar proportions of the two morphologies were observed. Melanocytic origin was evaluated using MelanA and HMB45 as melanoma markers. To calculate the percentage of Ki-67-positive cells (proliferation index), approximately five fields were captured in hotspot areas at 400x magnification and at least 500 marked and unmarked cells were analysed using Fiji/ImageJ software (National Institutes of Health) and the Cell Counter plug-in. Two independent evaluators conducted all analyses.

Mice with successful tumour formation were evaluated for metastatic dissemination. At the experimental endpoint, lungs, livers, and other organs exhibiting suspected macroscopic metastases were collected. One representative tissue section from each organ was analysed by H&E staining. When tumour-like cells were identified, sections were further stained with a human-specific Lamin A/C antibody to assess the presence of human cells. Additional confirmation was performed using melanoma-specific markers (HMB45 and MelanA). Metastatic spread was considered positive if detected in at least one mouse from the AM-PDX group.

### Patient survival analyses

For patients in clinical stages I–IV, Overall Survival (OS) and Disease-Specific Survival (DSS) were assessed based on the date of death. Only patients whose samples were freshly engrafted were included (n=39). Patients who underwent non-surgical treatment, lost to follow-up or reached the end of the observation period without experiencing the event were classified as “censored”. The percentage of survival was calculated as the cumulative number of censored participants up to each time point, divided by the number of patients at risk at the start of the study.

### Whole exome sequencing (WES) quality control and PDX filtering

A total of 119 samples were prepared for whole-exome paired-end sequencing at the Wellcome Sanger Institute (UK), comprising 48 tumours from 40 patients, 36 AM-PDX samples (including 20 from *in vivo* passage X1, 7 from X2, 6 from X3, and 3 from X4), and 35 blood samples. Samples were genotyped using Fluidigm CGP for pair checking before their library preparation. Exome capture was performed using the Agilent SureSelect Human All Exon V5 bait set. Libraries were prepared using Illumina standard protocol and sequenced on Illumina Novaseq 6000 (S4 Flowcell) for 100 bp paired-end runs, with an average of 190 million reads generated per sample. Reads were mapped to GRCh38 Human reference genome with bwa-mem v0.7.17^61^, PCR duplicates were marked with samtools markdup v1.12. Concordance between tumour – blood samples was assessed using Conpair v0.2^62^Samples were checked to have at least 80% of the baits covered with a minimum of 20X or more otherwise, they were excluded from further analyses unless specified. For AM-PDX samples, reads were additionally mapped to a customised mouse reference genome file referred here onwards as NOD_ShiLtJ_V1_PDX. This fasta file comprises all the chromosomes from the Nod/ShiLtJ v1 reference genome and GRCm39 Y chromosome sequences. Afterwards, mouse reads were removed using XenofilteR v1.6^63^. Plots and summaries of the total number and proportions of reads filtered were made using custom R scripts (See GitHub repository). After quality checks, 66 matched pairs (42 tumour-normal and 24 PDX-normal, respectively), and 17 unmatched samples (6 tumours and 11 PDX samples were available for analysis.

### Somatic variant identification and annotation

Somatic calling of single-nucleotide variants (SNVs), multi-nucleotide variants (MNVs), and short insertion and deletions (INDELs) was performed using cgpCaVEMan v1.15.2^64^, SmartPhase (v1.2.1)^65^ and cgpPindel (v3.10.0)^66^ respectively, with parameters and filters as described previously^67^Calling for matched tumour or PDX samples was performed using their respective buffy coat normal samples, and unmatched samples were called using a BAM file containing a 50X exome coverage of simulated 100bp paired end sequencing from the GRCh38 reference genome sequence without mismatches. All cgpCaVEMan and cgpPindel parameters for unmatched samples were identical to those used for matched samples, except for the estimated normal cell fraction in the tumour set to 0.

Variant consequence prediction was performed using The Ensembl Variant Effect Predictor (VEP) (v103)^68^. Given that a gene may have multiple transcripts, the canonical transcript – as defined in Ensembl – was used to determine the variant consequence. In addition, VEP was also used to include custom annotations from the COSMIC (v97)^69^, ClinVar (update 20230121)^70^, gnomAD (v3.1.2)^71^ and dbSNP (v155)^72^ databases. VCF files containing somatic mutations were combined, filtered, and converted to Mutation Annotation Format (MAF) using custom scripts for plotting using maftools v2.14.0 see Github. Differences in somatic variant types, SNVs, deletions and insertions, across tumour samples with PDX-X1 samples were evaluated using Pearson correlation function “cor.test”, to identify monotonic relationships.

### Somatic copy number calling and analysis

Copy number calling for tumour samples with matched normal controls was performed using the tool Sequenza v3.0.0 executed in Python3 v3.8.15 and R v4.2.1^73^. The R package copynumber v1.29.0.9 was used alongside Sequenza to incorporate the GRCh38 reference genome. BAM files from tumour/normal pairs served as input data for this analysis. For PDX samples, BAM files aligned to the GRCh38 reference genome and processed with XenofilteR (as described above) were used. Manual quality filtering of the output results was performed, and all samples with cellularity lower than 0.35 were excluded from further analysis.

Significant regions with frequent amplifications and deletions were identified using Gistic2 (v2.0.23) with default parameters and only one sample per patient^74^. Segmentation data and copy number estimates generated by Sequenza were used as input. Regions were considered significant when estimated q < 0.1. Focal and broad copy number scores per sample were obtained using the CNApp web application ^75^. CNApp was run with default parameters using Sequenza-derived segmentation data as input. Focal (FCS) and broad copy number (BCS) scores were compared between patient tumour and PDX-X1 samples, and across mutational status groups (4WT, *KIT*, *KRAS/NRAS/HRAS*, *NF1*) and hailstorm status (absent vs present). To identify monotonic relationships between tumour and PDX-X1 samples, Spearman’s rank correlation was performed using the “cor.test” function.

For tumour samples without matched normal controls, copy number calling was done using CNVkit (v0.9.10)^76^. Sex-specific reference profiles were generated using normal-against-normal copy number calling to filter out low-quality normal samples, following the protocols described by Chandramohan, R. et al. (2022)^77^. Copy number alterations were called using the batch subcommand, gains were defined as segments with a log₂ ratio > 0.585, and losses as segments with a log₂ ratio < - 0.4. To reduce noise-related artifacts, we filtered samples based on signal variability: we calculated the median absolute deviation (MAD) of log₂ ratios for each sample, normalized this value based on segment length, and applied a modified z-score threshold of 3.5 to exclude outlier samples.

To evaluate CNA patterns across patient and PDX samples, we used the “plotCNA” function in MesKit v1.8.0^78^. Copy number (CN) values from Sequenza were adjusted relative to the ploidy of each sample: Values of 0 were retained as 0, representing a deletion; values smaller than the ploidy but different from 0 were set to 1, representing a loss; values equal to the ploidy were set to 2, indicating neutrality; values greater than the ploidy, but not exceeding twice the ploidy, were set to 3, representing a gain; and values that were at least twice times the ploidy were set to 4, indicating an amplification.

### Identification of Hailstorms

Genomic hailstorm events were identified following the approach described by Wang, M. et al. (2024)^8^. Copy number transitions (CNTs) were defined as breakpoints separating neighbouring segments by a logR of 0.5 or more, adjusted for purity and ploidy. Chromosomal arms were classified as harbouring hailstorm aberrations if they met the following criteria: (1) a significantly increased CNT density compared to the genome-wide average (hypergeometric test, P < 0.0001); (2) the presence of at least one amplified segment exceeding threefold the background ploidy with a minimum size of 200 kb, and (3) overall width of at least 5 Mb between the left and right most CNTs of the aberration. For chromosomal arms larger than 50 Mb, a sliding window approach was applied (50 Mb window, 5 Mb step size) to improve detection sensitivity. However, variations in tumour purity and copy number segmentation performance can affect the accuracy of hailstorm evaluation. As a result, hailstorm regions present in multiple samples from the same patient may not be detected in some cases. To address this, we manually inspected all tumour samples from each patient or PDX, where at least one sample exhibited hailstorms.

### Construction of phylogenetic trees

Phylogenetic trees were constructed using MesKit^78^ v1.8.0 in R 4.2.2. Only somatic mutations were included, as well as samples that had tumour tissue with matched normal tissue. We combined the MAFs of human and PDX samples into a single file, keeping only patients for whom at least two samples were available. We then used the functions “getPhyloTree” to build and “plotPhyloTree” and plot phylogenetic trees for each patient with the maximum parsimony (MP) method and a minimum VAF of 0.06. We also compared the mutation data stored in the object generated by “getPhyloTree” with the data from COSMIC’s Cancer Gene Census (v97). Heatmaps showing public, shared, and private mutations were plotted with the function “mutHeatmap”.

### Genotyping

Genotyping was performed from blood samples using Infinium Multi-Ethnic AMR/AFR-8 v1.0 array (Illumina) at King’s College London and Infinium™ Global Screening Array (GSA) v3.0 (Illumina) at University College London UCL (UK). A total of 35 AM samples were successfully processed. Ancestry estimation was performed using the ADMIXTURE algorithm^37^. The 1000 Genomes Project super-population labels AFR (African), AMR (Admixed American), EAS (East Asian), EUR (European), and SAS (South Asian) were used as population references. The primary premise for this estimation is that an individual’s ancestry originates from distinct populations, providing an estimate of the proportion of the individual’s genome that is derived from each of these ancestral populations^79^.

### Fusion analysis

Gene fusions and their breakpoints were identified using RNAseq data of the tumours and AM-PDX samples with the software STAR-Fusion v1.11.11 using STAR v2.7.8a. The Trinity Cancer Transcriptome Analysis Toolkit genome reference library for GRCh38, which uses GENCODEv37 (Ensembl v103) gene annotations, was used. The results were annotated and validated in-silico using FusionInspector v2.7.0 with parameters: only fusions labelled as in-frame and out-of-frame, supported by ≥5 split reads and ≥5 spanning reads, with ‘Yes anchor support’ set to TRUE, left and right breakpoint entropy ≥1.5, and unknown coding effect or annotated as previously reported in normal tissues on the Genotype-Tissue Expression (GTEx) dataset were retained. Finally, the results were plotted using the Chimeraviz V1.26.082 package in R v4.3.1.

### scRNA-seq data processing and differential gene expression

The scRNA-seq data of 6 acral and 3 cutaneous primary samples were retrieved from GSE215121^46^. Raw sequencing data was processed using Cell Ranger v8.0.1, and the resulting count matrices from each sample were loaded into Seurat v5.3.0. Potential doublets were identified and removed using the scDblFinder v1.18.0 algorithm, and cells with high mitochondrial gene expression (>5%) were removed. To eliminate batch effects and harmonize data across samples, individual Seurat objects were merged and normalized using SCTransform. Dimensionality reduction was performed with Principal Component Analysis (PCA), and data integration was achieved using the Harmony algorithm, which aligns shared cell populations across AM and CM datasets while preserving biological differences. The identification of distinct cellular populations was performed using the shared nearest neighbour approach (FindNeighbors) and the Louvain algorithm (FindClusters) implemented in Seurat, which allows cells to be grouped into transcriptionally similar clusters. For low-dimensional visualization, Uniform manifold approximation and projection (UMAP) was used. Automated cell-type annotation was conducted using SingleR v2.6.0, which compares each cell’s transcriptome to reference datasets such as BlueprintEncode from the celldex v1.14.0 package. To identify differentially expressed genes that distinguish AM and CM, pairwise comparisons are performed using the FindMarkers function in Seurat, applying the MAST statistical test. The top 100 differentially expressed genes in the melanoma cluster were used to calculate a ratio based on the average expression of the AM-like and CM-like signatures. The relation between the gene expression level of *SF3B4* and AM/CM-like signatures was assessed using a linear model fitted and validated using stats package. Diagnostic plots confirmed the validity of the model, including the linearity of the relationship, normality of residuals, and homoscedasticity. All the analyses were performed in R v4.4.1.

### CRISPR screening

HEK293T cells were maintained in Iscove’s Modified Dulbecco’s Medium (IMDM) supplemented with 10% FBS and 1% penicillin-streptomycin. AM cell lines were cultured in RPMI 1640 10% FBS supplemented with 1% penicillin-streptomycin and 2mM L-glutamine. To generate Cas9-expressing cells, AM cells (AM007 and AM016) were transduced with lentivirus carrying the pKLV2-EF1a-Cas9Bsd-W plasmid (Addgene, 68343), previously packaged in HEK293T cells. Cells were selected with blasticidin (2.5 µg/mL), and Cas9 activity was assessed using a BFP/GFP reporter assay^42^. Activity was > 89% in both cell lines (data not shown).

The Human Improved Genome-wide Knockout CRISPR Library v1.1, a gift from Kosuke Yusa (Addgene, 67989), was packaged into lentivirus in HEK293T cells, and BFP expression was used for titration. Cas9-expressing AM cell lines were transduced with the CRISPR library in triplicate, using a multiplicity of infection (MOI) of 0.3, 8 μg/mL polybrene, and selected with puromycin (1.25 µg/mL for AM007 and 2.5 µg/mL for AM016). Cells were maintained throughout the screen at a minimum representation of 200X. Genomic DNA (gDNA) was extracted 14- and 21-days post-selection. The library was sequenced and used as a control.

Following initial demultiplexing, single-end sequencing reads were aligned to the Yusa genome-wide sgRNA library (90,709 sgRNAs) and quantified using pyCROQUET v1.5.1 (https://github.com/cancerit/pycroquet). Raw sgRNA counts per sample are provided in the GitHub repository. To account for variations in sequencing depth across samples, raw counts were normalised to a total of 45,354,500 sgRNA counts per sample (corresponding to 500x library representation). For each screen timepoint replicate, raw log_2_ fold-change (LFC) values were then calculated relative to the normalised baseline plasmid library counts. Specifically, for sgRNA *i*:

raw LFC*_i_* = log_2_ (normalised count *i*, screen +1) / log_2_ (normalised count *i*, library plasmid +1).

A pseudocount of 1 was added to both screen and plasmid library counts to prevent division by zero and to ensure defined LFC values for sgRNAs with zero normalised counts at a given replicate. To reduce noise from sgRNAs with insufficient representation, sgRNAs with a library-normalised count < 30 were removed.

To assess each screen timepoint’s ability to distinguish between non-essential and essential genes, we computed the Null-Normalized Mean Difference (NNMD) for each replicate^80^. The NNMD was calculated as the difference between the mean LFC of sgRNAs targeting reference pan-essential genes and the mean LFC of sgRNAs targeting reference non-essential genes, divided by the standard deviation of LFCs for sgRNAs targeting non-essential genes. Reference pan-essential (CEGv2) and non-essential (NEGv1) gene sets were obtained from the Hart Lab BAGEL resource (https://github.com/hart-lab/bagel). Raw LFCs were scaled such that the median LFC for sgRNAs targeting non-essential genes was set to 0, while the median LFC for sgRNAs targeting reference pan-essential genes was set to −1. To determine single-gene essentiality, scaled LFC values were input into BAGEL2^43^, which computes Bayes Factor (BF) scores per gene for each screen timepoint. Genes with BF > 10 were classified as essential.

To identify genetic dependencies specific to AM, we integrated data from our genome-wide CRISPR screens with results from the Avana Cas9 Library screens found in the DepMap 24Q4 release. Non-cancerous cell lines and engineered models were excluded from the analysis. The AM dataset included AM007, AM016, along with the three DepMap cell lines: WM3211 (DepMap ID: ACH-002508), M040416 (DepMap ID: ACH-002510), and MM160113 (DepMap ID: ACH-002512). The pan-cancer dataset comprised all cancer cell types, excluding all melanoma subtypes (cutaneous, uveal and vulval). BF scores were rank-normalised, and a Mann-Whitney U test was applied to identify significant gene vulnerabilities. Log fold changes were calculated using a code adapted from David Adam’s group (https://github.com/team113sanger/uveal_melanoma_specific_vulnerabilities/). Due to lack of statistical power to pass FDR correction, AM hits with p < 0.05 and LFC > 1.5 were considered as positive. Analysis was performed in R v4.4.3.

### Drug screening

Cells were seeded in 24-well plates at an optimized density for each cell line. Thirty compounds were diluted in DMSO in 3.16-fold steps to prepare a six-point dilution series, then further diluted 31.6-fold in RPMI 1640 with 10% FBS. Each concentration was added to cells in duplicate. The final DMSO concentration was 0.32% in all treated wells and 3.16% in untreated controls. Cell viability was measured by luminescence using an EnVision Multimode Reader (PerkinElmer) at baseline (24 hours after seeding) and 11 days after treatment start. Viability was normalized to DMSO controls. Nonlinear regression curves with a variable slope were generated using Prism (GraphPad, v9.0) to determine each compound and cell line’s IC50 (half-maximal inhibitory concentration). To visualise the pattern of drug screening response, the IC50 data were log-transformed, standardized, and analysed using customized heatmaps (ComplexHeatmap, circlize, and cluster packages). Analysis was done using R v4.2.3. The experiment was performed by Oncolines B. V. (https://www.oncolines.com/).

### RMC-7977 Sensitivity assays

Cells were seeded in 6-well plates at 0.75 to 1.5X10e5 cells per well and treated with different concentrations of RMC-7977 (Revolution Medicines and MedChemExpress) or vehicle control (DMSO). Plates were inserted into the Incucyte live cell imager for 6 days (144 hrs), and cell growth was analysed for percent plate coverage with IncuCyte Live Cell Analysis System. Nine pictures per well were taken every two hours. Treatment was renewed every 48-72 hrs.

For dose response curves, cells were seeded in 96-well plates (4-8X10e3 cells/well) and treated with RMC-7977 for 144 hrs. Treatment was renewed every 48-72 hrs. Cell viability was evaluated using the CellTiter Blue Assay (Promega). Fluorescence was measured at 560 nm and 600 nm using the SpectraMax ID3 plate reader (Molecular Devices). Viability was normalized to DMSO-treated wells, set to 100% viability, while wells lacking cells were set to 0% viability. To estimate the IC50, non-linear regression curves were calculated from the average values obtained using the Y=100/(1+10^((LogIC50-X) * HillSlope)) model and GraphPad Prism v10.0.

For both assays, at least three independent experiments with three technical replicates each were performed for each cell line.

### Western Blotting

Whole cell lysates were prepared and electrophoretically separated as described previously^81^. After electrophoresis by SDS-PAGE, membranes were incubated for approximately 16 hours with the following primary antibodies: anti-total-ERK (1:1000, Cell Signaling, 9102), phospho-ERK Thr202/Tyr204 (1:2000, Cell Signalling, 9106), anti-GAPDH (1:1000, Santa Cruz, sc-32233), anti-HSP90 (1:1000,Cell Signalling, 4877) diluted in 5% bovine serum albumin (BSA) in TBS-T 0.05% (0.12% Tris; 0.9% NaCl; 0.05% Tween-20). After washing, the membranes were incubated with secondary antibodies anti-rabbit IgG (1:10,000 - Cytiva, NA934) or anti-mouse IgG (1:10,000 - Thermo Fisher Scientific, 31430), both diluted in 5% milk in TBS-T for 1 hr. Proteins were detected using ECL SuperSignal West Pico Plus Chemiluminescent Substrate (Thermo Fisher Scientific) and visualized using a Chemidoc Imaging System (BIORAD). At least three independent experiments were performed.

### Statistical analysis

Analyses were performed using the statistical software GraphPad Prism v9.0 and 10. Data obtained were evaluated for normality using the Shapiro-Wilk test, and statistical tests were applied according to the data distribution. To compare two groups with non-parametric distribution, the Wilcoxon signed-rank test was used, while variables with parametric distribution were analysed using an unpaired T-test. Three or more groups were compared using one-way ANOVA followed by Tukey’s post hoc test. Benjamin-Hochberg FDR correction was applied when needed. Chi-square or Fisher test was used to evaluate the association between categorical variables. All *in vitro* experiments had at least three biological replicates for each cell line. All western blot experiments were run on samples coming from at least three separate biological experiments. Survival rates were analysed using the Kaplan–Meier method and log-rank test. All comparisons when p < 0.05 were considered statistically significant.

## Data Availability

Sequencing data is available at the European Genome-Phenome Archive (EGA)

## SUPPLEMENTARY METHODS

### Mutational and copy number signature analysis

Mutational signatures were analyzed using the tool SigProfilerExtractor v1.1.23. For Single-base substitution mutational signature analysis, SNVs and INDELs that passed the previously described filters were used as input data and signatures were extracted using the 96 SBS context with default parameters for exome data. For copy number signature analysis, we used Sequenza segmentation data as input and the following parameters: minimum_signatures = 1, maximum_signatures = 10, and nmf_replicates = 100, while keeping the remaining settings as default (optimized for WES data).

### Association Analyses

The categorical variable hailstorm status for each chromosomal region, using the Wilcoxon signed-rank test following a post-hoc method (Benjamin-Hochberg test) for multiple testing correction, using function “wilcox.test”. To evaluate the association between categorical variables and hailstorm status of chromosomal regions 11q, 22q, and 5p (absent vs present), Fisher’s exact test was performed using the function “fisher.test”. This set of analysis was performed in R v4.4.1. Engraftment rate (PDX vs no PDX) was evaluated across clinical variables such as tumour site, sex, age, anatomic site, Breslow depth, mitotic index, ulceration, and necrosis. This set of analyses was performed using GraphPad Prism v9.

### Transcriptome mapping, filtering, quantification and quality control

RNA from 60 samples (41 tumours and 19 PDXs) was used for stranded RNAseq tagged library preparation with standard Oligo dT pulldown protocols. Subsequently, sample libraries were multiplexed and sequenced using Illumina Novaseq 6000 (S4 Flowcell) at the Wellcome Sanger Institute (UK). Sequencing data aligned to the GRCh38 human reference genome using STAR v2.5.0c. For PDX samples, reads were mapped to the custom mouse reference genome NOD_ShiLtJ_V1_PDX (generated as mentioned above) alongside a custom GTF file that contained the ENSEMBL v107 annotation for the Nod/ShiLtJ v1 reference genome and GRCm39 Y chromosome using STAR v2.7.10. Mouse reads were identified and excluded from BAMs using XenofilteR v1.6. Cleaned human-aligned AM-PDX and tumour BAM files were used to assess expression using htseq-count v0.13.5 with the parameters “-m union --stranded reverse” and ENSEMBLv103 annotation. Samples with < 20 million read pairs were removed from subsequent analysis. Finally, counts were transformed to transcripts per million (TPM) and used for correlation analyses. For more detailed information, see GitHub repository.

### Cell death assay

Cells (1×10e5) were seeded in 24-well plates in duplicate and treated with 50nM of RMC-7977 for 48 hours. Cells and supernatants were centrifuged and washed. Pellets were incubated in PBS + 2% FBS for 30 min followed by staining with Annexin V-APC (Invitrogen) for 15 min, and 7AAD for 5-15 minutes (eBioscience). Stained cells were diluted in Annexin V-APC Apoptosis buffer 1x (eBioscience). The experimental controls included unstained cells, cells treated with 50% ethanol (as a positive control for 7AAD staining), and cells treated with DMSO. Annexin V-APC (633nm) and 7AAD (488nm) stained cells were analyzed using BD FACSCanto II (BD Biosciences). In total 10.000 events were analyzed per measurement. Data was analyzed using FlowJo™ v10 Software (BD Life Sciences). Three independent experiments with technical duplicates were performed.

### Sanger Sequencing

Genomic DNA (gDNA) was used to amplify a 223 bp fragment of exon 15 covering the coding region of codon 600 of the *BRAF* gene. The amplification was performed using the primers 5’-CATAATGCTTGCTCTGATAGG-3’ (forward) and 5’-GGCCAAAAATTTAATCAGTGG-3’ (reverse) with the AccuPrime® Taq DNA Polymerase (Thermo Fisher Scientific) on a Veriti® 96-Well Fast Thermal Cycler. The cycling protocol started with 95°C for 2 minutes, followed by 35 cycles of denaturation at 95°C for 15 seconds, annealing at 55°C for 30 seconds, and extension at 68°C for 30 seconds. The resulting PCR product was purified using the GFX PCR DNA and Gel Band Purification kit (Merck). *BRAF*^V600E^-mutant D10 melanoma cell line was used as a positive control.

---

**EXTENDED DATA FIGURE 1 (related to Fig. 1):**
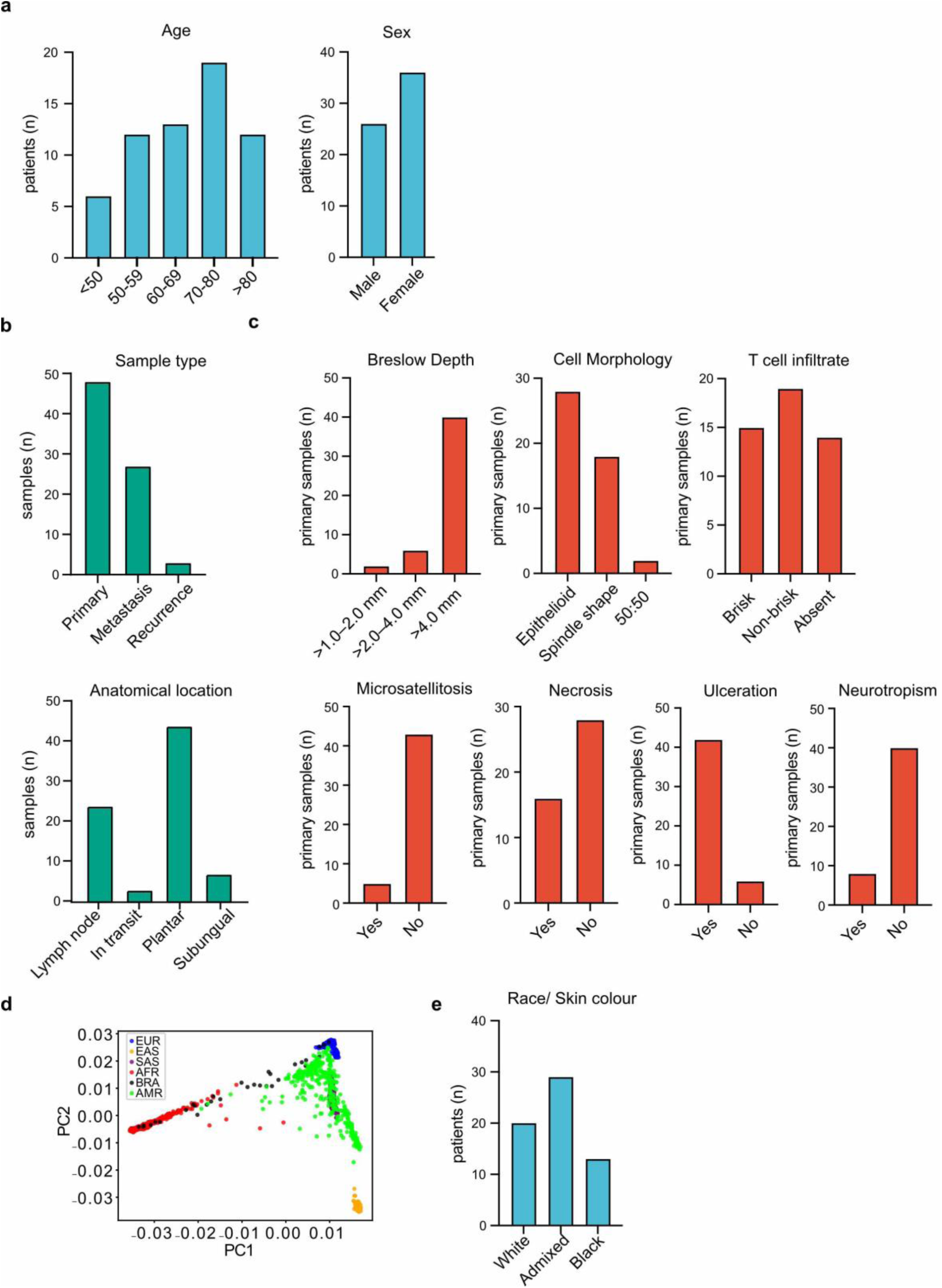
A) Number of patients according to age and biological sex (n=62). B) Number of samples according to sample type and anatomical location (n=78). C) Number of primary tumour samples according to histopathological characteristics (n=48). D) Plot depicting the first two principal components identified using genotyping data of the study population (BRA, black) and five 1000 Genome superpopulations: Europeans (EUR, blue), East Asians (EAS, orange), South Asians (SAS, purple), Africans (AFR, red), and Amerindians (AMR, green). E) Number of patients according to self-reported skin colour (n=62).

**EXTENDED DATA FIGURE 2 (related to Fig. 2):**
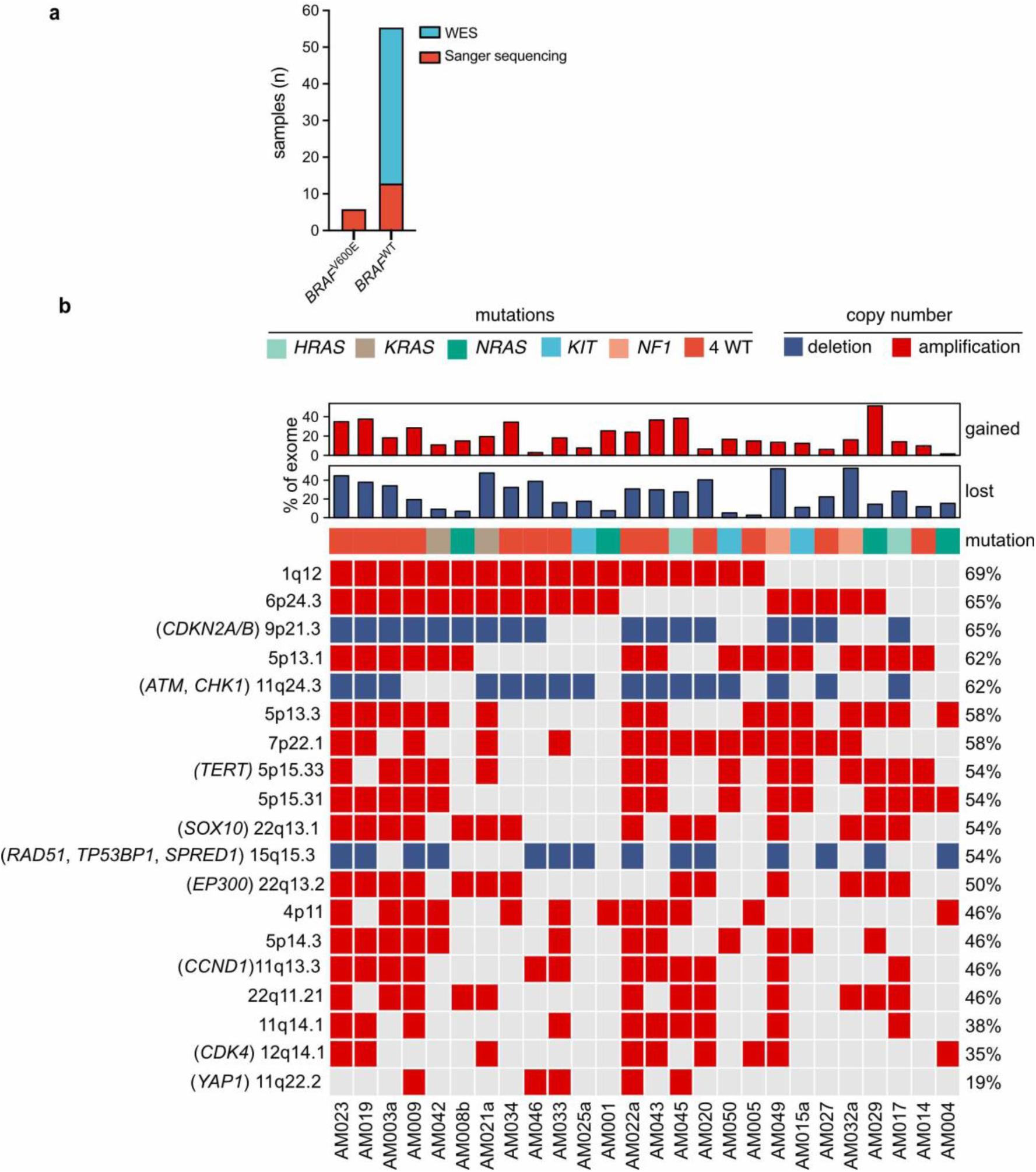
A) Bar graph illustrating the frequency of patient tumours with and without *BRAF*^V600E^ mutations assessed by whole exome sequencing (blue) or Sanger sequencing (red) (n=78). B) Distribution of copy number alterations (rows) per sample (columns) according to SEQUENZA AND GISTIC2. Regions with significant amplifications (red) and deletions (blue) and cytoband where the altered chromosomal segment was detected are shown. The presence of mutations in *KRAS/NRAS/HRAS*, *KIT* or *NF1*, as well as quadruple wild-type tumours (4WT) are also illustrated. Bars on top represent the percentage of the whole exome exhibiting copy number gains or losses. Frequencies (right) were calculated from 26 tumours, one sample per patient.

**EXTENDED DATA FIGURE 3 (related to Fig. 2):**
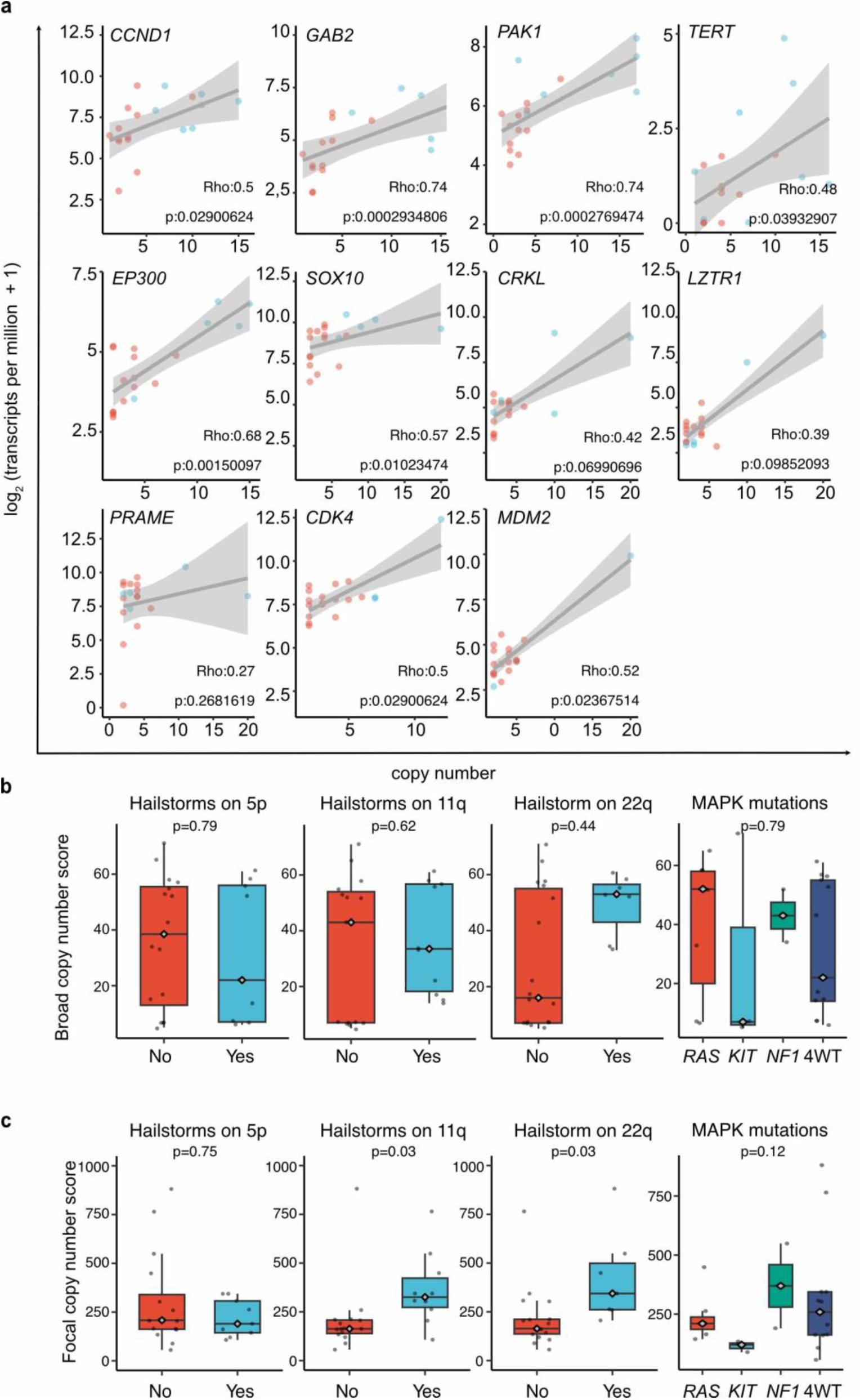
A) Correlation of copy number gains and gene expression of genes detected in hailstorms. Blue and red dots represent samples with and without hailstorms, respectively. B, C) Comparison of broad (BCS) and focal copy number (FCS) burden across tumours with and without hailstorm events on chromosome arms 11q, 22q and 5p and across *RAS*, *KIT*, *NF1*, and quadruple wild-type (4WT) tumours. For hailstorm comparisons, Wilcoxon tests were used, and p values were adjusted for multiple testing using the Benjamin– Hochberg. For mutation comparisons, Kruskal−Wallis tests were used. Box plot features: rhombus symbol = median; box limits = upper and lower quartiles; whiskers = up to 1.5 × interquartile range; points = outliers.

**EXTENDED DATA FIGURE 4 (related to Fig. 2):**
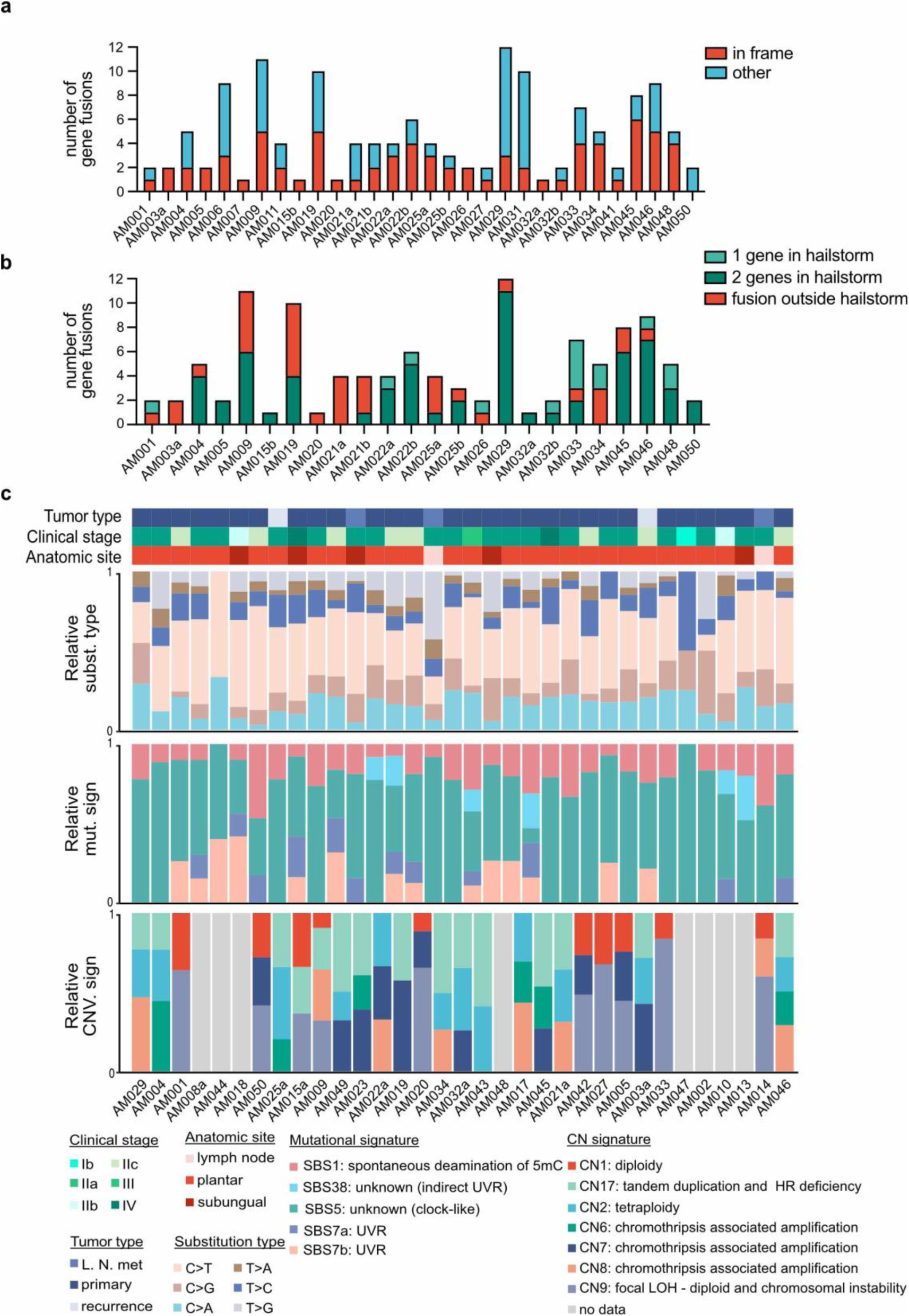
A) Number of gene fusions in patient tumours detected in the RNAseq data (n=30). In-frame gene fusions are illustrated in red. B) Number of gene fusions according to their location. Outside (red) and within (green) hailstorm regions. Green tones indicate if one or both genes were detected within the limits of the hailstorm regions (n=24). C) Proportions of relative substitution type, mutational signatures for single base substitutions and for copy-number aberrations are shown in stacked bars per patient tumour sample. Clinical stage, tumour type and anatomic site are shown for each sample.

**EXTENDED DATA FIGURE 5 (related to Fig. 3):**
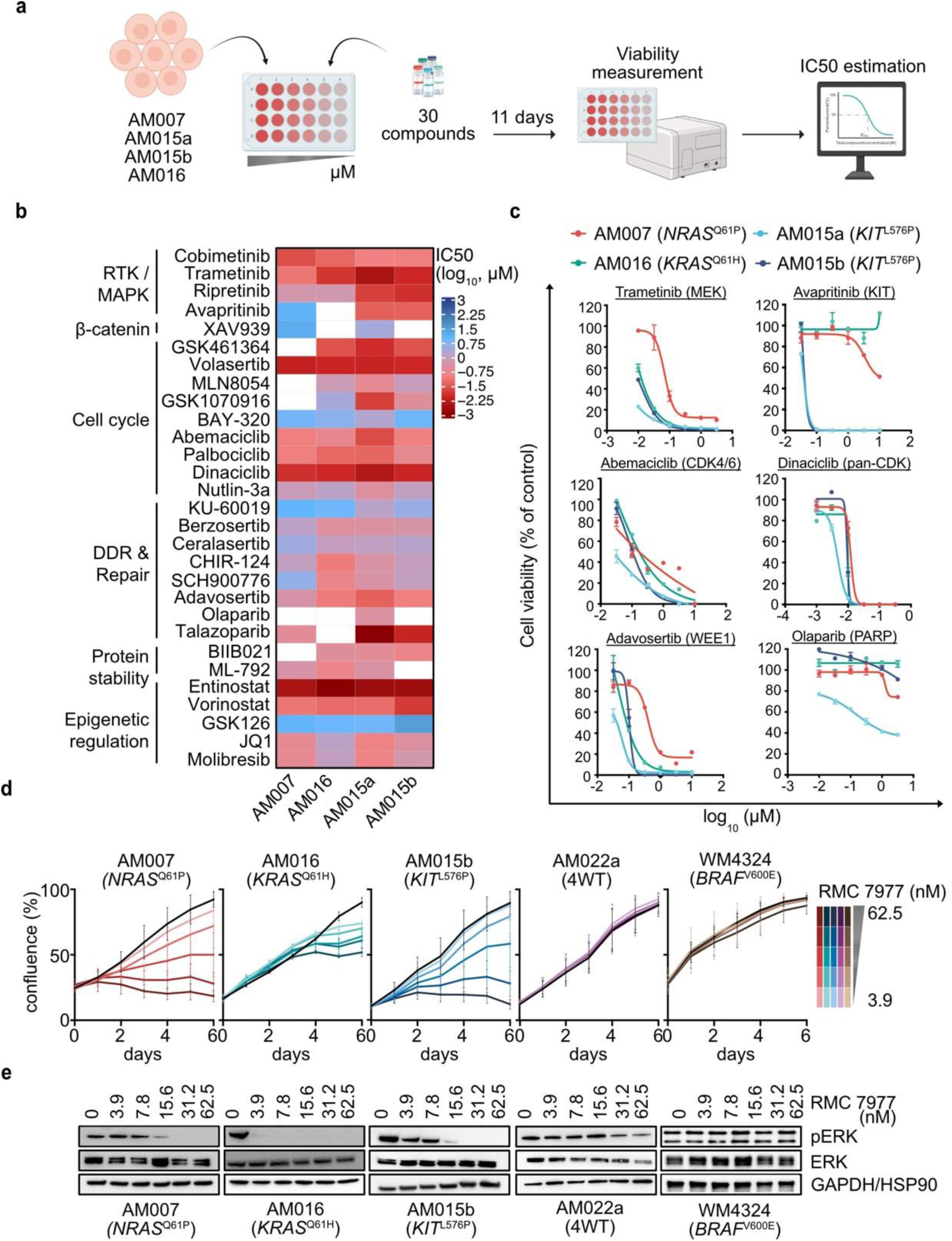
A) Proportions of successful tumour growth and generation of AM-PDX (green) and no tumour growth (red) for samples (n=73). Samples were separated according to sample processing in fresh (n=54) and frozen (n=19). Three samples were excluded from the analysis due to not meeting the criteria for evaluating tumour take. Success rate is indicated as % on top of the columns. Fisher’s exact test. B) Time from sample inoculation to palpable tumours (200mm^3^). Each dot represents a sample (dark blue: fresh, light blue: frozen). Centre line = median. C) Histopathological comparison of patient tumours and corresponding AM-PDX models. Slides were stained for Haematoxylin-Eosin (H&E), Ki-67 (proliferation marker) and Melan-A (melanoma marker). Four representative models are shown. Scale bar 50 µm. D) Comparison of the proliferation indexes of patient tumours and their corresponding AM-PDX as calculated by the percentage of Ki-67-positive cells in each sample (p=0.977, Paired t test). E) Bar graphs showing the percentage of mouse reads detected on the AM-PDXs RNAseq data using the Xenofilter tool. The remaining reads were derived from human RNA, corresponding to the xenografted patient tumour.

**EXTENDED DATA FIGURE 6 (related to Fig. 3):**
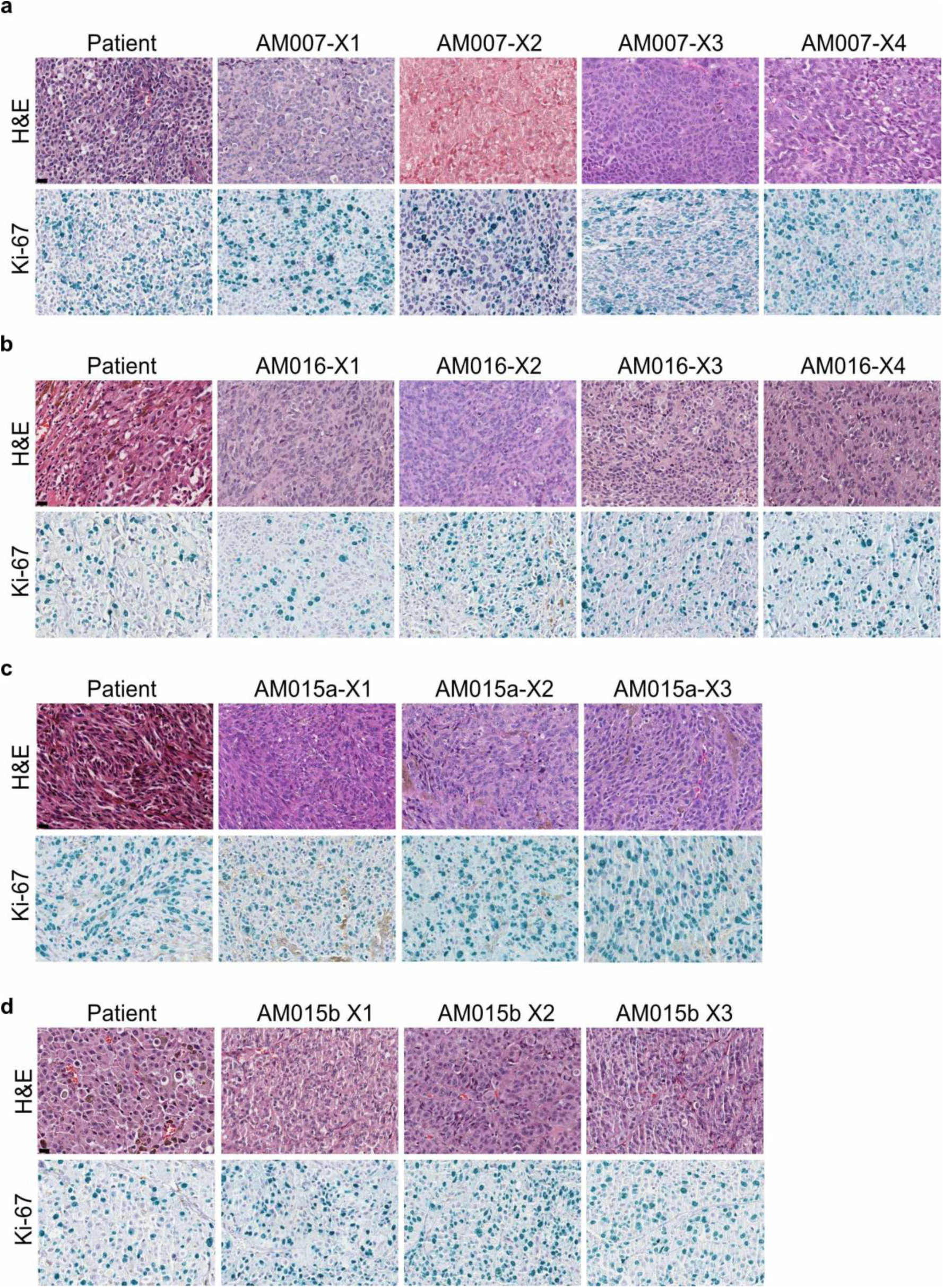
Histopathological characteristics of AM-PDX models across subsequent *in vivo* passaging. Four representative models are shown. Sections of patient tumours and AM-PDXs were analysed by H&E (upper panel) and Ki-67 (bottom panel) staining. A) AM007. B) AM016. C) AM015a D) AM015b. Scale bar 20 µm.

**EXTENDED DATA FIGURE 7 (related to Fig. 4):**
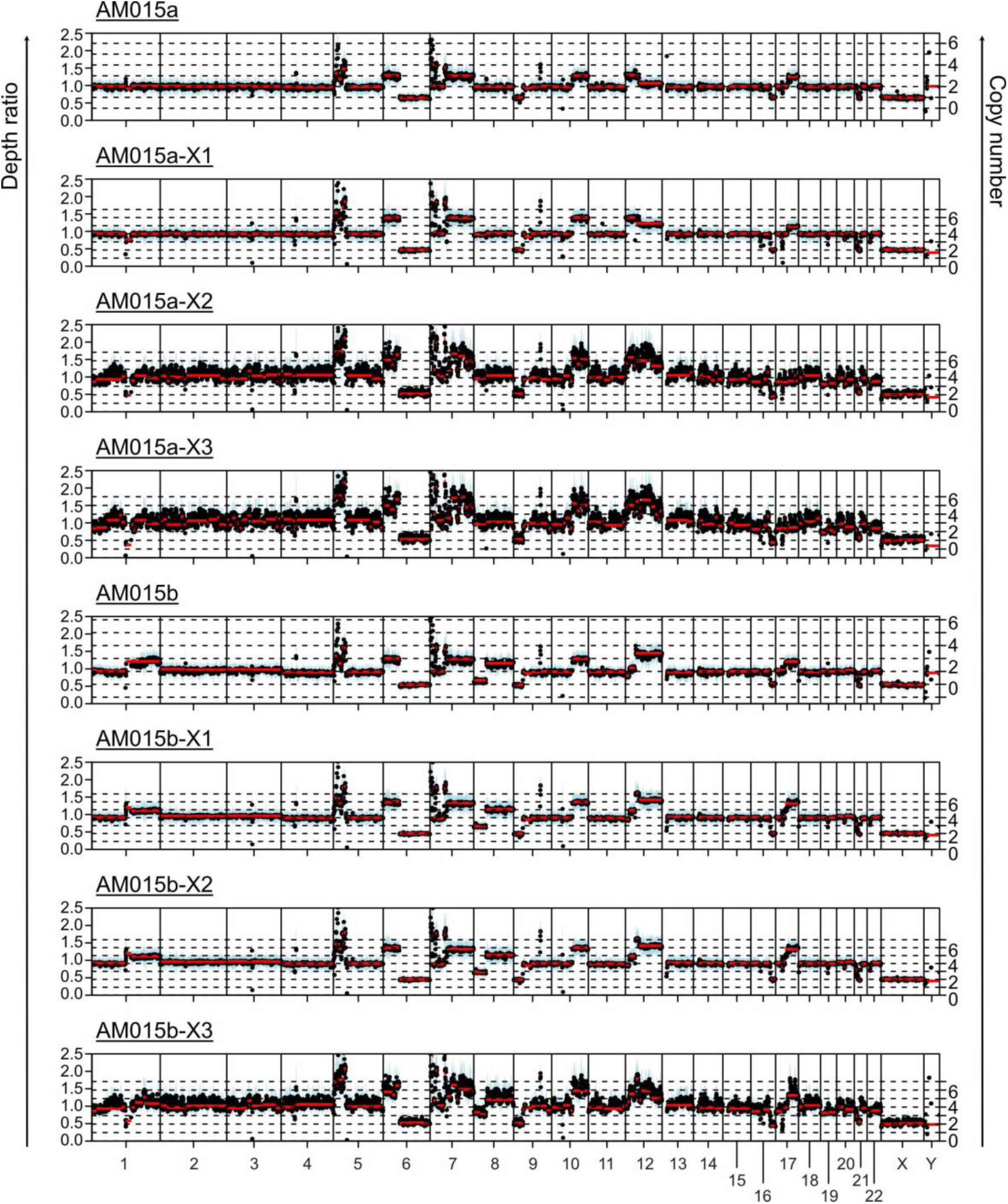
Representative example of hailstorm events observed in patient tumours (AM015a: primary tumour; AM015b: lymph node metastases) and three passages of their corresponding AM-PDX models. The left Y-axis and black dots represent depth ratios (tumour/normal) across different genomic regions. Blue shading represents variability in the depth ratio estimation. The right Y-axis and red lines show the estimated copy number for each chromosome segment.

**EXTENDED DATA FIGURE 8 (related to Fig. 4):**
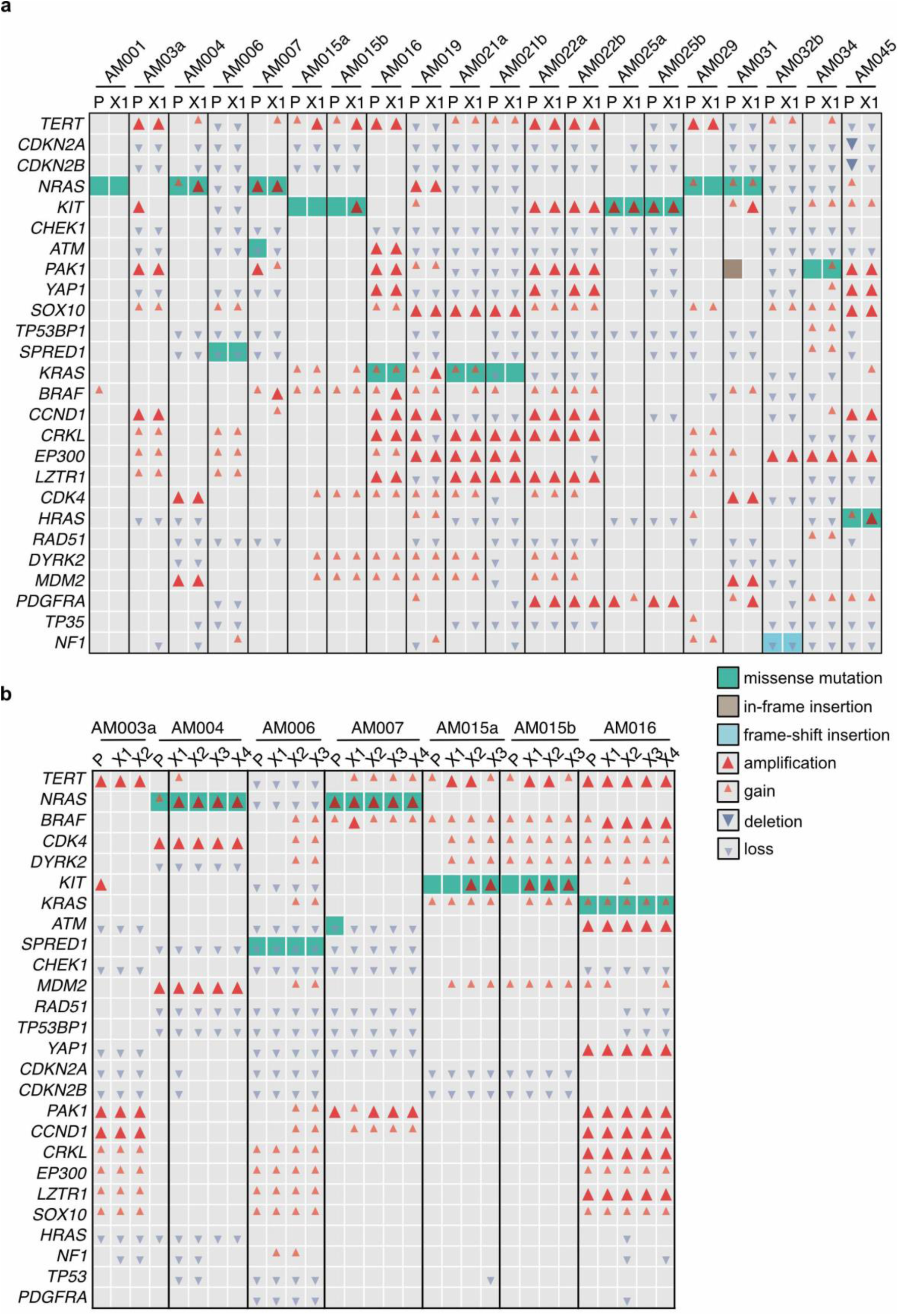
Genes commonly altered in patient and AM-PDX samples. A) Patient tumours and X1 passages are shown for 20 AM-PDX models. B) Patient tumours and different passages are shown for representative AM-PDX models. Genes catalogued by the COSMIC or previously related to melanoma are illustrated Type of alterations are illustrated with different colours or symbols. cn: copy number; Amplification: cn ≥ 4; Gain: 2 < cn < 4; Loss: 0 < cn < 2; Deletion: cn = 0.

**EXTENDED DATA FIGURE 9 (related to Fig. 4):**
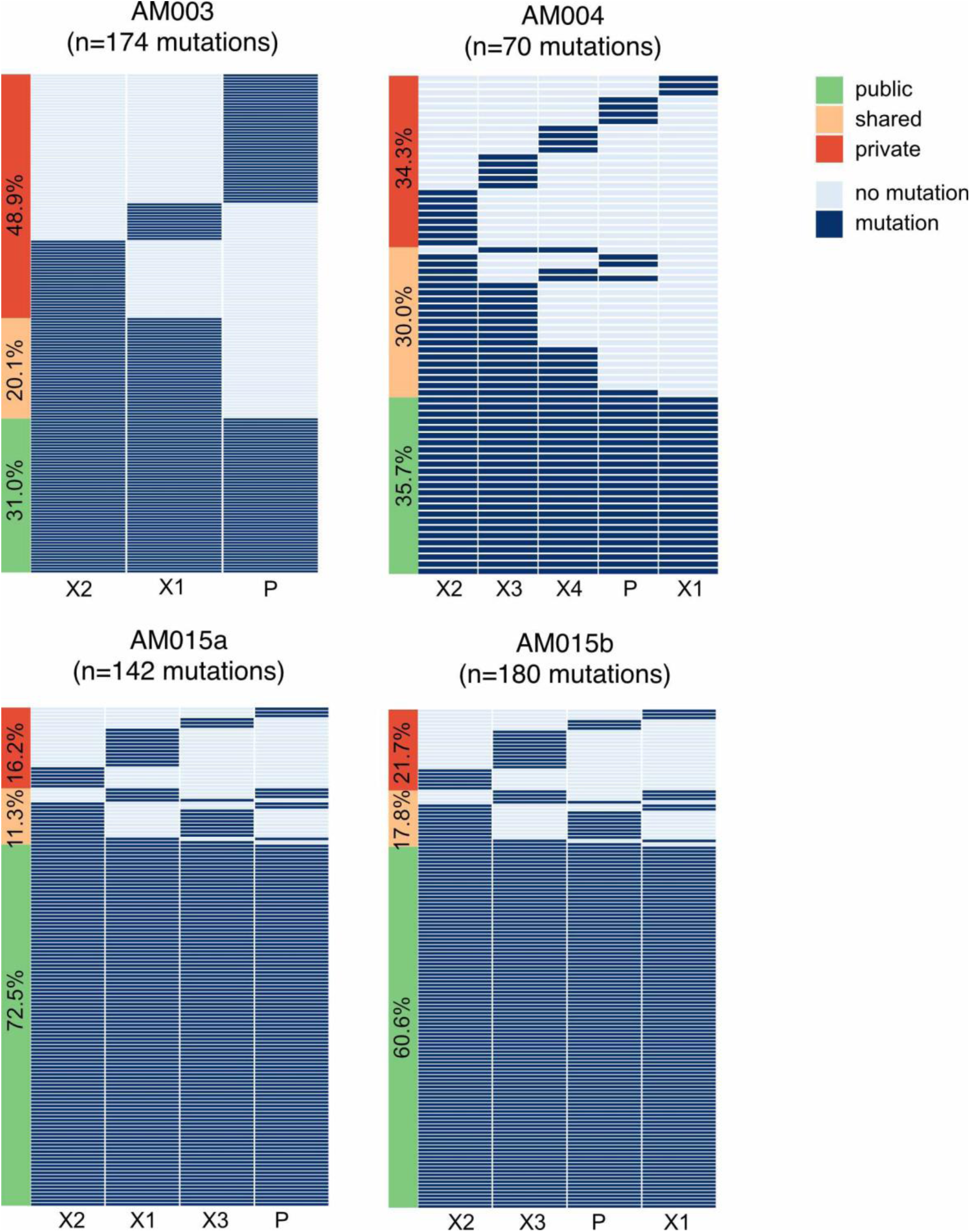
Heatmap illustrating the frequency (%) of somatic SNV and indels alterations in patient tumours and the corresponding AM-PDX models across passaging. Total number of mutations detected is shown. Public (detected in all samples) shared (detected in more than one sample) and private (detected in only one sample) are shown.

**EXTENDED DATA FIGURE 10 (related to Fig. 5):**
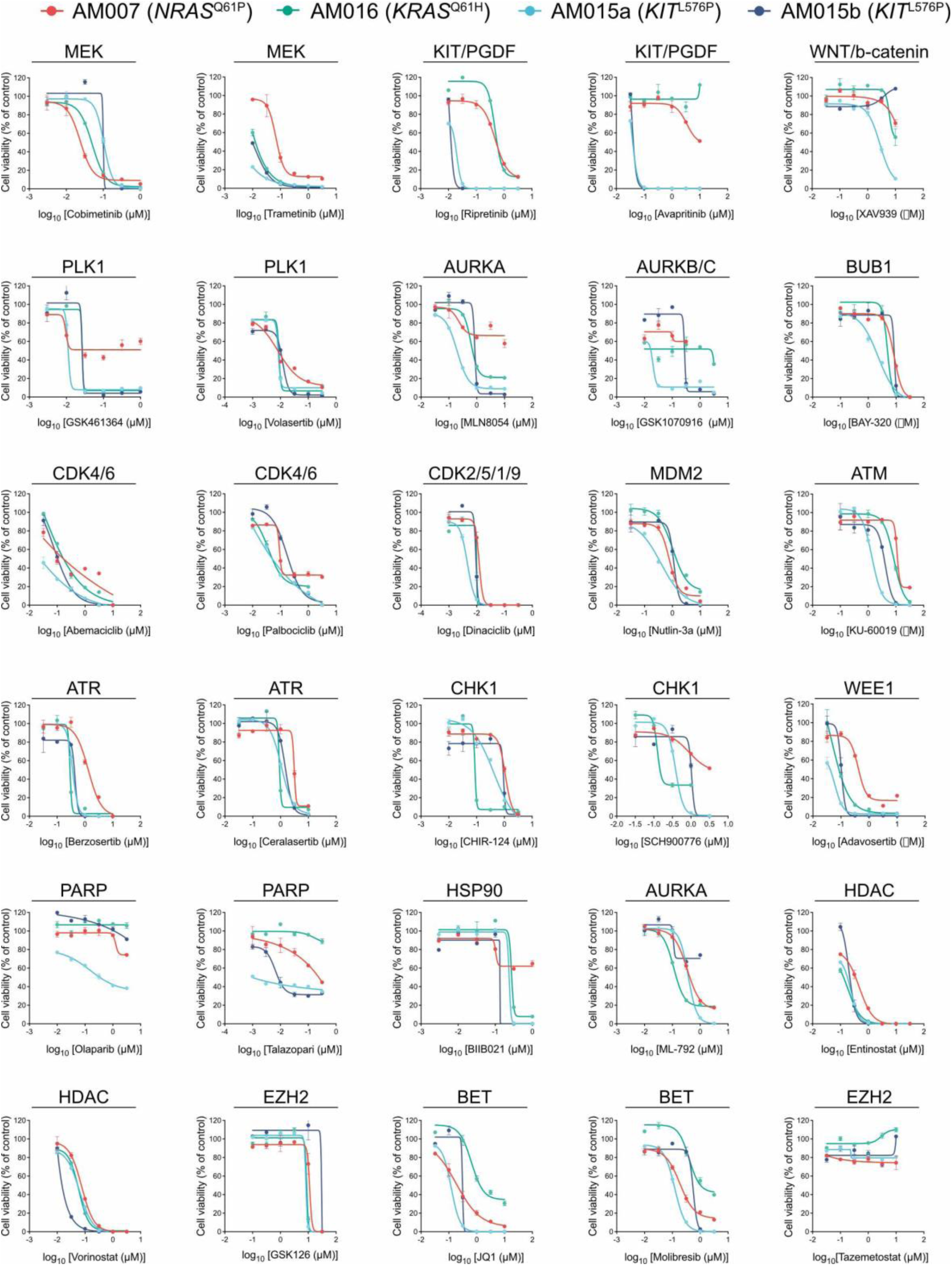
A) Cells were treated with a panel of 30 small-molecule inhibitors for 11 days at different concentrations. Cell viability was measured using ATPlite and IC50 values were calculated. Dose response curves are shown for all drugs tested.

**EXTENDED DATA FIGURE 11 (related to Fig. 5 and 6):**
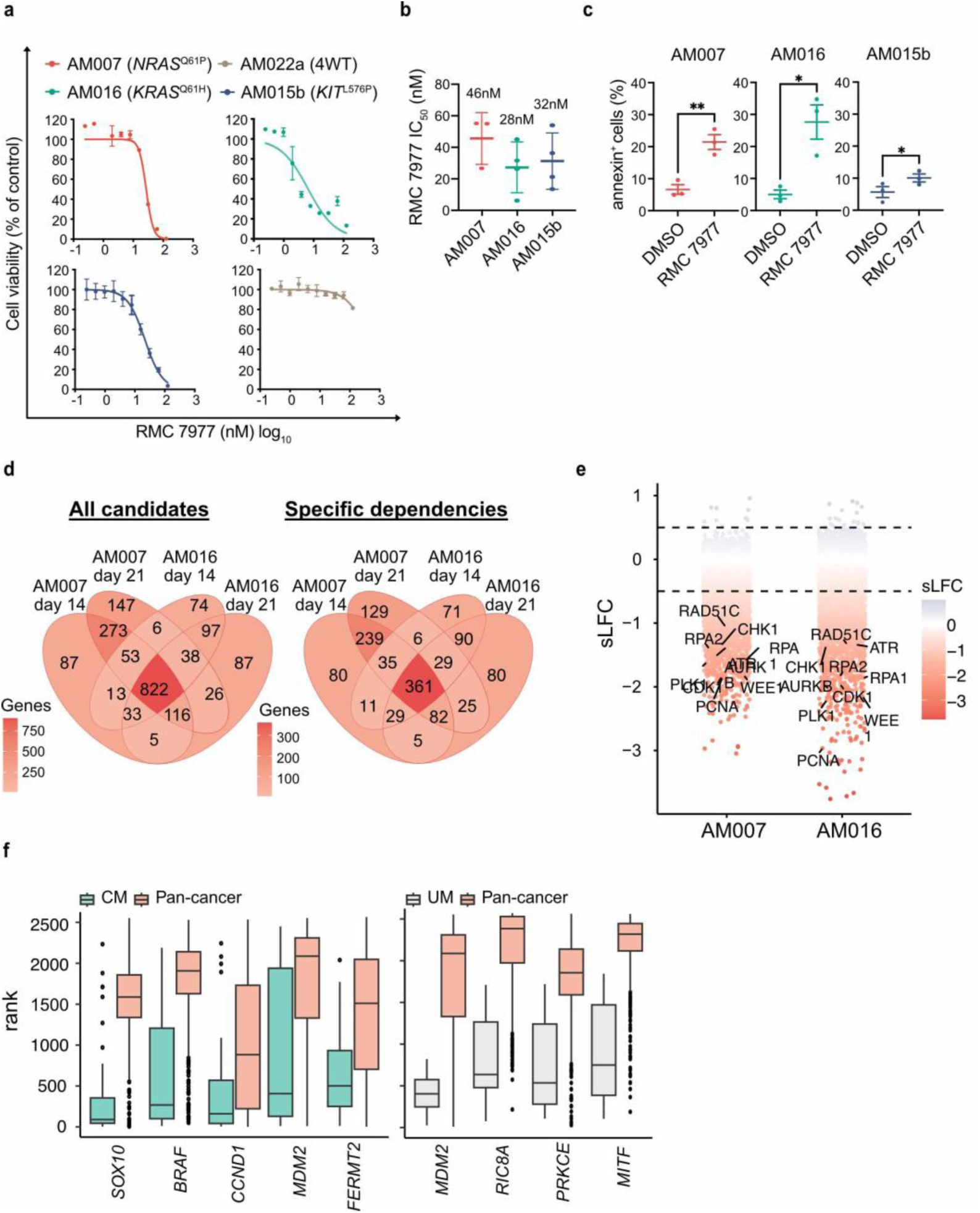
A) Cells were treated with DMSO or increasing concentrations of RMC-7977 for six days. Treatment was renewed after three days. Cell viability was measured using Cell titer blue. The percentage of viable cells is depicted as mean (+/− SD) on the Y-axis. Log_10_-transformed IC50 values are shown on the X-axis. A representative experiment for each cell line is shown. B) IC50s derived from dose response curves. n=3 biological replicates with 3 technical replicates each. Data is shown as mean +/− SEM and mean value is shown on top. C) Flow cytometry analysis of AM cell lines treated with 50 μM RMC-7977 for 48hs. Percentage of Annexin V+ staining compared to DMSO is shown as mean (+/− SEM) (One-tailed paired Student’s t test, *: p < 0.05 and ** p < 0.01). D) Venn diagram depicting the overlap of genes identified as known essential genes (top, n=822) and specific dependencies (bottom, n=361) according to BAGEL analysis across different conditions. The central overlap highlights genes consistently identified in both cell lines and time points. E) Waterfall plot illustrating common dependencies ranked by their scaled Log Fold Change (sLFC). Selected genes are shown. Dotted lines indicate significant thresholds (0.05 < sLFC < −0.05). F) Genes identified as essential in cutaneous (CM, n=56 cell lines) and uveal melanoma (UM, n=10 cell lines) compared to other tumour types (pan-cancer, n= 1010 cell lines) using DepMap datasets (p=0.05; Mann-Whitney U test). Only genes identified as more essential in CM or UM are shown. Box plot features: centre line = median; box limits = upper and lower quartiles; whiskers = 1.5× interquartile range; points = outliers.

## SUPPLEMENTARY TABLE LEGENDS

**Supplementary Table 1:** Clinical characteristics of patients included in the study cohort

**Supplementary Table 2:** Histopathological characteristics of AM samples

**Supplementary Table 3:** Mutations detected in AM samples, one sample per patient

**Supplementary Table 4:** Hailstorms detected in patient tumours and AM-PDXs

**Supplementary Table 5:** Gene fusions detected in patient tumours and AM-PDXs

**Supplementary Table 6:** Mutations detected after in vivo passaging of AM-PDXs

**Supplementary Table 7:** Main driver mutations and copy number alterations detected in patient tumours and AM-PDXs

**Supplementary Table 8:** Main driver mutations and copy number alterations detected in AM-PDXs from which cell lines were derived

**Supplementary Table 9**: Small molecule inhibitors included in the Pharmacological screen along with their respective targets and dose range used

**Supplementary Table 10:** List of vulnerabilities shared between AM007 and AM016 cell lines

**Supplementary Table 11:** Top 100 differentially expressed genes between melanoma cells in AM and CM subtypes from scRNAseq datasets

## Acknowledgements

We would like to thank the patients and their families for agreeing to participate in this study and for providing access to their samples and clinical data. We are thankful to the Program of Immunology and Tumour Biology for technical, administrative, and scientific support. We also thank Ana Frazao, Paulo César Moraes, Diego Gomes, and Luciana Morreuw from the National Tumour Bank, Nicole Scherer from the Bioinformatics Core Facility, and the teams from the Division of Human Pathology and the Animal Experimentation Facility at INCA. We are grateful to Dr. Nataliya Shifrin and Revolution Medicines Inc. for providing RMC-7977. Finally, we thank Tim Bishop for insightful discussions. This work was financially supported by the Medical Research Council (MR/S01473X/1, 2019) to D.J.A., P.A.P., and C.D.R.-E.; the Melanoma Research Alliance Pilot Awards (825924) to C.D.R.-E., D.J.A., and P.A.P. and (1264411) to A.E.A., C.D.R.-E., and P.A.P.; Science Without Borders - CNPq (401116/2014-0) to P.A.P.; the CNPq International Program, including fellowships to A.C.M.S.S and F.C.A (442091/2023-0); FAPERJ (E-26/010.002427/2019,E-26/010.002508/2016, E-260003/015681/2021) to P.A.P.; the Royal Society Newton Advanced Fellowship (NAF\R2\202163) to P.A.P.; the MRC Dermatlas Project (MR/V000292/1) to D.J.A.; NIH grants (U54 CA224070, SPORE P50 CA174523, R01 CA259295, P30 CA010815) to M.H, and to the Dr. Miriam and Sheldon G. Adelson Medical Research Foundation. Additional support was provided by the Wellcome Sanger Institute International Fellowship (to P.A.P. and C.D.R.-E.), the CNPq Productivity Fellowship (309661/2023-4) to P.A.P., and the Wellcome Trust through a Career Development Award (227228/Z/23/Z to C.D.R.-E). Students and trainees at INCA were supported by fellowships from the Ministry of Health. This research was funded in part by the Wellcome Trust 220540/Z/20/A. For the purpose of Open Access, the author has applied a CC BY public copyright licence to any Author Accepted Manuscript version arising from this submission.

## Author Contributions

P.A.P., S.S.B., F.C.A., R.M.C.N., M.M.P., and D.G.C. collected and processed biological samples; S.S.B., R.M.C.N., F.C.A., and M.M.P. generated the PDX models and cell lines; P.A.P, J.P.S., and D.G.C performed nucleic acid extractions; A.R.B.N. performed histopathological review of samples assisted by S.S.B., J.P.S, R.M.C.N., and P.A.P.; P.V.F. performed immunohistochemistry; J.L.O, L.F.N., and A.C.M. assessed patients and provided access to clinical data; J.L.O. performed patient surgeries and provided biological samples; M.M.P., J.P.S, J.L.O., and P.A.P collected and reviewed clinical data; A.C.M.S.S., A.C.F., M.C.V.H, R.C.W.R., P.B.S, J.B., P.S.R.B., J.B., and I.S.W. performed bioinformatic and statistical analyses; F.C.A., R.O.L., L.S.A.S.M., and R.F. conducted and analysed the CRISPR screens; F.C.A. and D.G.C. conducted and analysed drug screens; A.C.M.S.S., R.M.C.N., and G.A. performed the RMC-7977 experiments supervised by A.E.A; I.V. coordinated the development of coding/GitHub repositories; A.C.M.S.S. and P.A.P supervised and interpreted analyses. A.C.M.S.S. and P.A.P wrote the manuscript and designed the figures with support from S.S.B., F.C.A., C.D.R.E., and D.J.A; This study was jointly supervised by C.D.R.E., D.J.A., and P.A.P with critical input and support from M.H. and A.E.A. All authors read, reviewed, and approved the final manuscript.

## Data and code availability

Sequencing data is available at the European Genome-Phenome Archive (EGA) under the following accession numbers: EGAS00001005663 (WES), EGAS00001008150 (RNAseq), EGAS00001008231 (Xenofiltered WES), EGAS00001008232 (Xenofiltered RNAseq), and EGAS00001008230 (CRISPR KO). Public single-cell datasets were retrieved from GSE215121^46^. The remaining data are available in Supplementary Information or Supplementary data files. All code used in the analyses is available at GitHub (https://github.com/team113sanger/Acral_Melanoma_PDX_models_from_Latin_Americ a).

## Competing Interests

The authors declare no competing interests.

## Notes

### Competing Interest Statement

The authors have declared no competing interest.

### Author Declarations

This study was conducted at the Brazilian Natioal Cancer Institute after approval by the Institutional (CEP) and National (CONEP) Research Ethics Committees (CAAE 98752818.6.0000.5274) including patient recruitment and informed consent, the generation of patient-derived xenografts biorepository, collection of demographic, clinical and histopathological data and molecular and functional analyses.

### Summary of Updates

The order of the authors in the website was not correct. The oder in the pdf file was correct. The only revision was updating the order on the website to match the pdf.

